# The epidemiology of neuropathic pain: an analysis of prevalence and associated factors in UK Biobank

**DOI:** 10.1101/2022.07.26.22278063

**Authors:** Georgios Baskozos, Harry L Hébert, Mathilde M V Pascal, Andreas C. Themistocleous, Gary J Macfarlane, David Wynick, David L H Bennett, Blair H Smith

## Abstract

Previous epidemiological studies of neuropathic pain have reported a range of prevalences and factors associated with the disorder. This study aimed to verify these characteristics in a large UK cohort. A cross sectional analysis was conducted of 148,828 UK Biobank participants who completed a detailed questionnaire on chronic pain. The *Douleur Neuropathique en Quatre Questions* (DN4) was used to distinguish between neuropathic pain (NeuP) and non-neuropathic pain (Non-NeuP) in participants with pain of at least 3 months’ duration. Participants were also identified with less than 3 months’ pain or without pain (NoCP). Binomial and multinomial regression were used to identify factors associated with NeuP compared to Non-NeuP and NoCP respectively. Chronic pain was present in 76,095 participants (51.1%). The overall prevalence of NeuP was 9.2% (13,744/148,828). NeuP was significantly associated with worse health-related quality of life, having a manual or personal service type occupation and younger age compared to NoCP. As expected NeuP was associated with diabetes and neuropathy, but also other pains (pelvic, post-surgical and migraine) and musculoskeletal disorders (rheumatoid arthritis, osteoarthritis and fibromyalgia). Additionally, NeuP was associated with pain in the limbs and greater pain intensity and higher BMI compared to Non-NeuP. Female gender was associated with NeuP when compared to NoCP, whilst male gender was associated with NeuP when compared to Non-NeuP. This is the largest epidemiological study of neuropathic pain to date. The results confirm that the disorder is common in the general population and is associated with a higher health impact than non-neuropathic pain.

## 1. Introduction

Chronic pain, usually defined as pain lasting for three months or more [53], is the leading cause of disability worldwide, when all pain conditions are taken into account, including low back pain, headache disorders and neck pain [23]. Neuropathic pain (NeuP) is a particularly severe form of chronic pain, arising as a direct consequence of a lesion or disease effecting the somatosensory nervous system [52]. Examples of common causes of NeuP include diabetes, HIV and chemotherapy treatment for cancer (causing painful peripheral neuropathies), herpes zoster (causing postherpetic neuralgia), multiple sclerosis, surgery, stroke and spinal cord injury [12]. NeuP is likely to be a major contributor to the global burden of chronic pain [7].

From a clinician’s perspective, it is important to distinguish NeuP from other forms of pain, for example nociceptive pain, which arises from actual or threatened damage to non-neural peripheral tissue. NeuP is generally unresponsive to analgesics such as non-steroidal anti-inflammatory drugs (NSAIDs) or opioids. Rather gabapentinoids (gabapentin and pregabalin), tricyclic antidepressants (TCAs) and serotonin-norepinephrine reuptake inhibitors (SNRIs; duloxetine and venlafaxine) are recommended as first- and second-line treatments [18]. Nonetheless, these medications for NeuP provide greater than 50% pain relief in less than half of people treated. Furthermore, analgesics in general, particularly opioids and gabapentinoids, can potentially cause harm, providing an even greater emphasis on appropriate use [26].

In order to identify NeuP in the community and the general population, screening tools such as the *Douleur Neuropathique en Quatre Questions* (DN4) [8], Self-Administered Leeds Assessment of Neuropathic Symptoms and Signs (S-LANSS) [5] and the painDETECT [20] have been developed. These rely on typical symptoms experienced in NeuP including burning, electric shocks, pins and needles and tingling. This has allowed epidemiological studies to be conducted. Estimates of NeuP in the general population suggest the prevalence is 7-10% [28], increasing to around 20-30% in people with diabetes [1,2,10,13]. Previous studies have also reported greater prevalence of NeuP, as with chronic pain overall, in older people, women and people from areas of high social deprivation [43].

However, due to differences in screening tools used and sample selection biases between studies, there is a large amount of heterogeneity in findings. For example, in Hong Kong the reported prevalence of NeuP was 5.1% using the Identification Pain Questionnaire [60], whereas in Kuwait it was 17.8% using the DN4 [62]. Furthermore, whilst a Canadian study reported a significantly increased risk of NeuP in males [54], other studies have reported an increased risk in females [9,50]. Reliable information on NeuP prevalence and associated factors in the general population are vital for developing prevention and management strategies. The UK Biobank has recently re-phenotyped participants for chronic pain, creating, to the best of our knowledge, the largest available cohort for the study of pain, including NeuP. The size and scale of this cohort provides an ideal platform with which to validate the findings of previous studies and identify novel associations.

The aim of this study was therefore to describe the epidemiology and clinical characteristics of NeuP in the UK Biobank.

## 2. Methods

This study is reported using the Strengthening the Reporting of Observational Studies in Epidemiology (STROBE) guidelines for cross-sectional studies (Supplementary Table 1).

### 2.1. Dataset

#### 2.1.1. The UK Biobank cohort

UK Biobank comprises 501,518 volunteers recruited between 2006-10 (at ages 40-69 years) from across Great Britain. It consists of demographic and health data as well as linked bio-samples and specialist investigations such as imaging in subgroups [46]. A brief assessment of Chronic pain was completed by all participants at first assessment (http://biobank.ndph.ox.ac.uk/showcase/field.cgi?id=6159) though it did not include validated questionnaires enabling categorisation of chronic pain and differentiation of NeuP from non-neuropathic pain (non-NeuP). The authors BS, DB, DW and GJM participated in the design of a set of revised UK Biobank pain questionnaires [6] between 2017 and 2018. This was tailored to UK Biobank and based on validated questionnaires already in routine use and as far as possible aligned with established international consortia studying chronic pain such as Generation Scotland [41,42] and DOLORisk [27,40]. These were designed to focus on the most prevalent causes of chronic pain and associated comorbidities and risk factors, including: musculoskeletal pain, NeuP and headache. The choice of items to include for NeuP was based on international recommendations made for phenotyping in genetic studies [29].

#### 2.1.2. Re-phenotyping for neuropathic pain

Once approved by the UK Biobank scientific panel, a successful pilot test of 10,000 participants was undertaken in April 2019. The chronic pain phenotyping survey was then sent to all currently active UK Biobank participants in May 2019 who had consented to further electronic contact and had an active email address (n=335,587). This has been completed by 167,203 individuals.

The revised follow up UK Biobank pain questionnaires first asked about medical history, based on a list of conditions that commonly lead to chronic pain. These included musculoskeletal conditions such as osteoarthritis and rheumatoid arthritis but also important aetiological factors for NeuP such as diabetes, nerve damage, neuropathy and post-herpetic neuralgia.

Participants were asked to indicate if they had been suffering from pain for more than three months (defined as chronic pain [53]); if yes, they were asked if they experienced pain all over the body and if they answered yes they were asked to report pain intensity on average over the previous 24 hours on an 11-point visual analogue scale (VAS; 0 being no pain, 10 the worst possible pain) and directed to part one of the ACR questionnaire for preliminary diagnostic criteria for fibromyalgia [59]. If they did not experience pain all over the body they were directed to a list of body sites, where they indicated all the locations in which they had experienced pain in the previous three months. They also rated the pain intensity at each location over the last 24 hours, on an 11-point visual analogue scale (VAS; 0 being no pain, 10 the worst possible pain). They then identified which of these pains had bothered them most during the previous three months. As many people with chronic pain have multiple pain conditions, the subsequent pain-related questionnaires asked the participants to complete them with regard to their most bothersome pain. To assess the quality of their pain, participants completed the self-reported items on the *Douleur Neuropathique en Quatre Questions* (DN4) questionnaire which is a validated tool for NeuP screening based on pain quality [8]. The characteristics of the most bothersome pain were captured with bespoke questions based on the Brief Pain Inventory (BPI) – Short Form [11], asking about pain intensity over the previous 24 hours and on average. Participants were asked to rate the relief they obtained from pain medication, and how their pain impacted different areas of their lives (e.g. general activity, mood, sleep).

The EQ-5D-5L[30] was used to ask about health-related quality of life in more detail; this included a 0-100 VAS, where the participants were asked to select the number that best represented their health at that time, and an assessment of the extent of limitations experienced in different domains of life (mobility, self-care, usual activities, pain or discomfort, and anxiety and depression).

The presence of peripheral neuropathy in the legs and feet was assessed with the self-complete component of the Michigan Neuropathy Screening Instrument (MNSI) [16], for those participants who answered yes to the presence of at least one of the following in the medical history section: cancer pain, diabetes or nerve damage other than diabetic neuropathy. These conditions are common causes of length-dependent peripheral neuropathy, and symptoms are expected to be present distally. Finally, participants were asked to complete psychosocial questionnaires about depression (Patient Health Questionnaire-9, PHQ-9 [35]) and fatigue (Fatigue Severity Scale, FSS [36]).

Body Mass Index (BMI) was calculated from height and weight data measured during the initial Assessment Centre visit in 2006-2010. Ethnic background, job and current employment status were also collected during the initial Assessment Centre visit using the UK Biobank touchscreen questionnaire. Age of participants at the time of completion of the pain phenotyping questionnaires was calculated from the year of birth, collected at recruitemnet, and the time the questionnaires were completed. Index of Multiple Deprivation was calculated using distinct domain of deprivations that can be recognised and measured separately and were collected during the Initial Assessment visit. Main diagnosis was obtained from the summary field of the distinct primary diagnosis, coded using the International Classification of Diseases (ICD-10), which were recorded from the participants hospital inpatient record during 2013-2022. Full background information about the follow-up UK Biobank pain questionnaire, and the questions included is available at https://biobank.ctsu.ox.ac.uk/crystal/ukb/docs/pain_questionnaire.pdf.

For analysing this study, we divided the cohort into three groups:

1. Chronic neuropathic pain (NeuP): participants who reported having pain for more than three months AND who scored 3 or more on the DN4, N = 13,744
2. Chronic non-neuropathic pain (Non-NeuP): participants who reported having pain for more than three months AND who scored less than 3 on the DN4, N = 62,351
3. No chronic pain (NoCP): participants who did not report having pain, OR whose reported pain had lasted less than three months, N = 72,733 Only fully completed DN4 questionnaires were considered for group allocation. Chronic pain participants with an incomplete DN4 were not classified.

### 2.2. Statistical analysis

Data was collected electronically by completion of a structured online questionnaire. As non-white ethnic backgrounds were rare in UK Biobank (2.2%), we grouped together all Black, Asian and Minority ethnicities (BAME) into one group. Of the total UK Biobank cohort, 1.6% were BAME, 0.6% were mixed ethnicity and the remaining 97.8% were white.

Occupations were encoded according to the Departments of National Statistics Standard Occupational Classification (SOC 2000) major groups. The Index of Multiple Deprivation, a weighted composite score showing relative deprivation, i.e. the higher the score the more deprived the individual, was used as a measure of poverty.

In order to determine differences between groups we carried out omnibus tests: one-way ANOVAs for normally distributed data or the Kruskal Wallis test for non-parametric data. These were followed up with Tukey’s HSD post-hoc tests. Associations between groups and categorical variables were tested using the Chi-Square test. Significant associations were followed up by pairwise Bonferroni corrected chi-square post-hoc tests, the standardised residuals showing higher or lower than expected ratios and the associated p.values are shown in supplementary figures. Univariable tests were followed by multivariable modelling with binomial or multinomial, multiple logit regression depending on the number of levels of the dependent variable. In multivariable modelling, missing values were imputed in 20 cycles of multiple imputations by chained equations using the predictive mean matching algorithm. Before model fitting, we removed variables with >30% missing values, variables with low variance, the least informative variable (i.e. lower variance) from highly correlated pairs (Pearson’s correlation coefficient >0.8) or those directly measuring pain. Missing value percentages for all variables are in supplementary table 2. In multivariable modelling we used the outcome as the dependent variable and a set of uncorrelated independent variables that included the self-reported quality of life measured by the EQ5D index, sex, age, ethnic background, job, index of multiple deprivations, the 3 first principal components of genetic variation and binary tags for self-reported diseases: Diabetes, Other Neuropathy, Fibromyalgia, Chronic Fatigue Syndrome, Osteoarthritis, Rheumatoid arthritis, Cancer pain, Carpal Tunnel Syndrome, Complex Regional Pain Syndrome, Chronic post-surgical pain, Gout, Migraine, Pelvic pain. The genetic variation principal component loadings were downloaded from UK Biobank and were calculated by PCA analysis on 101284 SNPs.

Model coefficients were aggregated across imputations using Rubin’s rules. For the significant model coefficients (one-way ANOVA p.value <0.05), we present exponentiated aggregate model coefficient estimations and the associated 95% confidence intervals.

## 3. Results

### 3.1. Demographics and socio-economic status

167,203 participants completed the pain experience phenotyping (response rate 49.8%). There was an over-representation of participants who were female, of younger age, lower BMI and less socially deprived in the group that chose to complete the pain re-phenotyping questionnaire compared to the rest of the UK Biobank cohort who did not complete the pain phenotyping, regardless of whether they were sent a questionnaire, supplementary table 3. 148,828 out of the 167,203 (89%) participants had no chronic pain or chronic pain with a fully completed DN4 questionnaire, which can be used as a screening tool for NeuP (see methods). The participants with completed DN4 had the same deprivation levels as those that had not completed the questionnaire, but there were more females, of slightly higher age and higher BMI than the participants with incomplete DN4, supplementary table 4. 13,744 participants reported chronic NeuP; 62,351 participants reported chronic Non-NeuP; 72,733 participants reported no chronic pain. The prevalence of NeuP in this cross-sectional cohort was therefore 9.2% (13734/148828). Age and sex were significantly associated with outcome (p.value < 0.001): participants with NeuP were younger than those with Non-NeuP (p.value <0.01, means difference −0.33, 95%CI[-0.5, -0.16]), figure 1A and supplementary figure 1A. Participants who reported chronic pain, both Neuropathic (NeuP) and Non-Neuropathic (Non-NeuP), were more likely to be female than those participants without chronic pain (59% vs 52%, p.value < 0.001), figure 1B and supplementary figure 1B. Ethnic background was significantly associated with outcome (p.value < 0.001), figure 1C. 90.7% of participants with NeuP were white British; in the total UK Biobank cohort 97.8% of participants were white British. There was a higher proportion of whites in the Non-NeuP (98.34%) vs the other groups, while BAME were more likely to report NoCP and NeuP (1.58%) versus Non-NeuP (1.19%),, supplementary figure 1C. BMI was significantly associated with pain grouping (p.value <0.001): BMI was significantly higher in participants with NeuP vs Non-NeuP (p.value <0.01, means difference 1.14, 95%CI[1.04, 1.23]) and in participants with Non-NeuP vs NoCP (p.value <0.01, means difference 1.86, 95%CI[1.76, 1.96]), supplementary figure 1D. Both employment status and occupation/job type were significantly associated with the outcome (p.value < 0.001). People with NeuP were more likely to report that they were “Unable to work because of sickness or disability” compared to those with Non-NeuP and NoCP, figure 2A and supplementary figure 2A. People with NeuP were more likely to work in “Elementary occupations”- simple and routine tasks, involving the use of hand-held tools and in some cases considerable physical effort; “Process, plant and machine operatives” - operate and monitor large scale, and often highly automated, industrial machinery and equipment; and “Personal service occupations” compared with the other two groups, figure 2B and supplementary figure 2B. The multiple deprivation index was significantly associated with the outcome (p.value < 0.001) and was higher (consistent with increased deprivation), in people with NeuP vs Non-NeuP (p.value <0.01, means difference 2.04, 95%CI[1.76, 2.32]) and in NeuP vs NoCP (p.value <0.01, means difference 2.34, 95%CI[2.06, 2.61]). It was also slightly higher in and Non-NeuP vs NoCP (p.value < 0.01, means difference 0.3, 95%CI[0.14, 0.46]), supplementary figure 2C.

**Figure 1.**
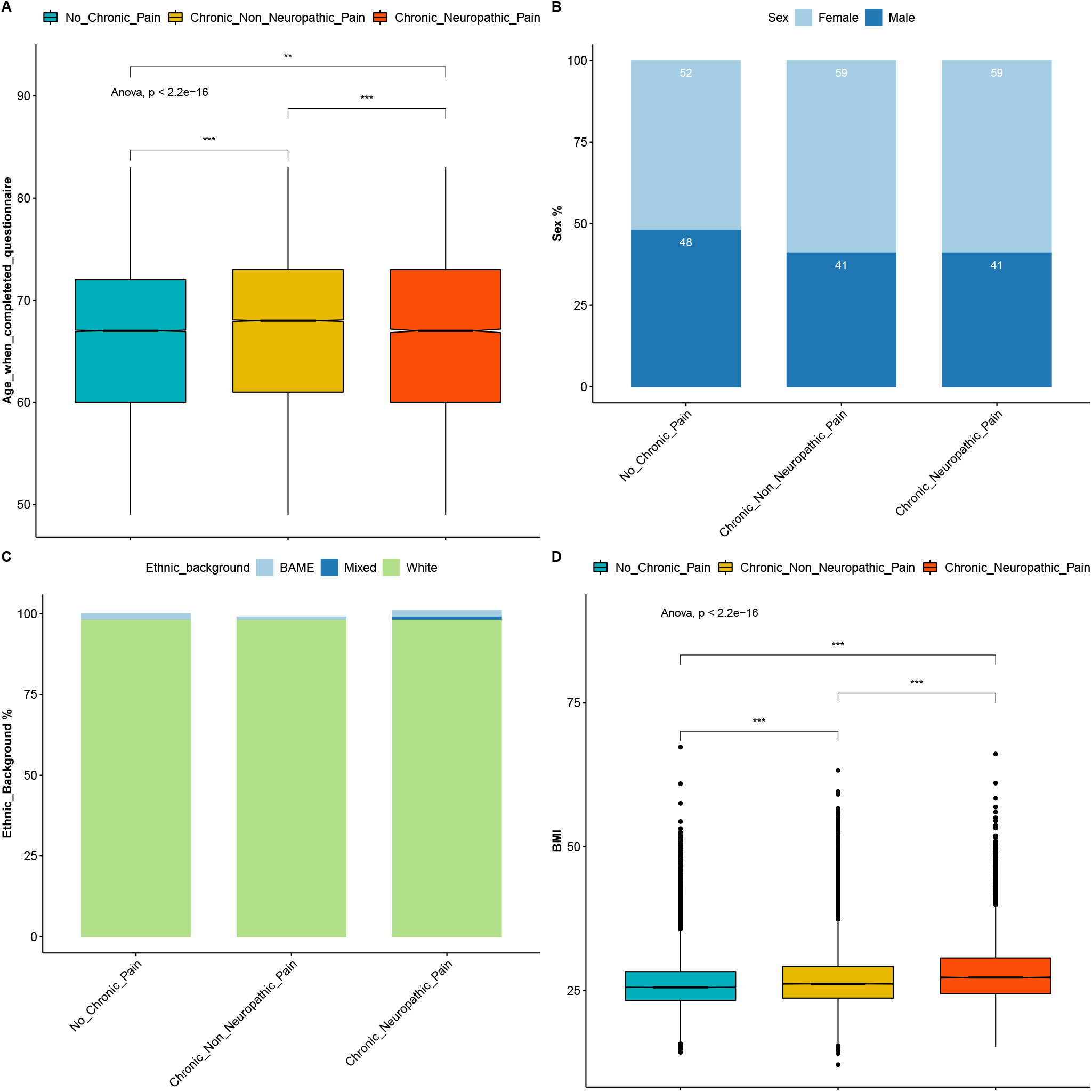
Demographics of participants with chronic NeuP, Non-Neup and NoCP. A: Boxplots show the age of participants when questionnaires were completed for each group. Notched line represents the median. B: Stacked bar plots show the sex distribution across the three groups. C: Stacked bar plots show the ethnic background distribution across the three groups. D: Boxplots show the BMI of participants across the three groups. Omnibus ANOVAs are labelled in the plot and are followed-up by t-tests between groups. P.value < 0.001 is coded as ***, p.value < 0.01 is coded as ** 0.01, p.value < 0.05 is coded as *. Post-hoc tests are shown in supplementary figure 1.

**Figure 2.**
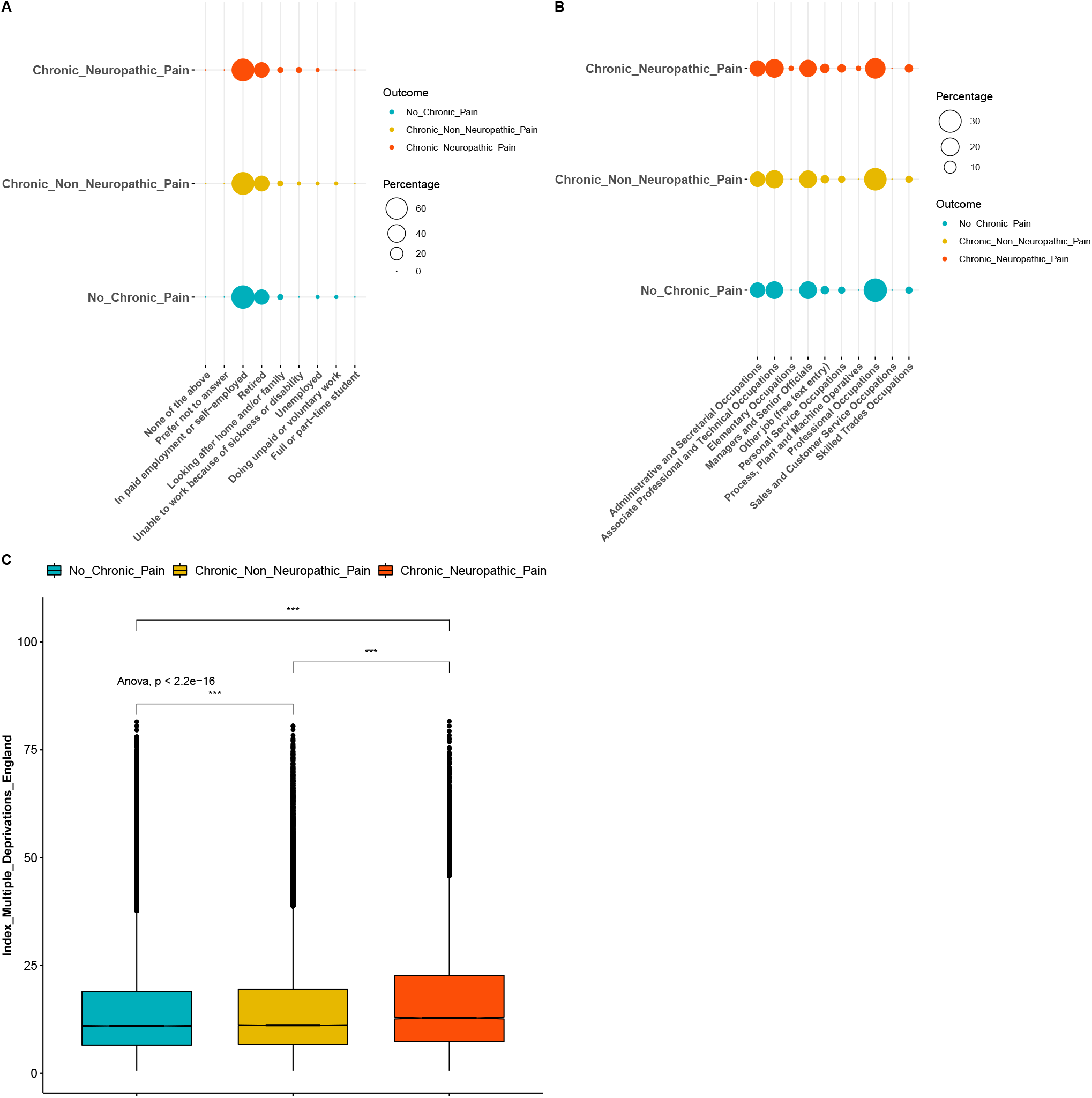
Socio-economic status of participants with chronic NeuP, Non-Neup and NoCP. A: Dotplot shows the employment status of participants for each group. The group is colour coded, the respective percentage is coded in the size of the dot. B: Dotplot shows the occupation of participants for each group. C: Boxplots show the index of multiple deprivations across the thee groups. Omnibus ANOVAs are labelled In the plot and are followed-up by t-tests between groups. P.value < 0.001 is coded as ***, p.value < 0.01 is coded as ** 0.01, p.value < 0.05 is coded as *. Post-hoc tests are shown in supplementary figure 2.

### 3.2. Pain rating and quality of life

People with NeuP reported higher pain severity ratings for the self-reported most bothersome pain (p.value <0.001), figure 3A. The most bothersome pain in people with NeuP was reported to be in the feet, hands or legs was more frequently; whereas reporting the most bothersome pain was reported to be in the back, hip, knee, stomach, abdomen or head most frequently by people with Non-NeuP, figure 3B. The quality of life as measured by the EQ5D normalised index was dependent on outcome (p.value < 0.001) and was significantly lower (p.value <0.01) in people with NeuP vs NoCP (means difference −0.22, 95%CI[-0.23, -0.22]) and Non-NeuP vs NoCP (means difference −0.14, 95%CI[-0.14, -0.14]), figure 4A and supplementary figure 3. The association between NeuP and reduced quality of life was consistent across all body locations figure 4B. People with NeuP were more likely to have “Diseases of the nervous system” than by both the other groups and “Certain infectious and parasitic diseases” than people with NoCP, figure 5 and supplementary figure 4.

**Figure 3.**
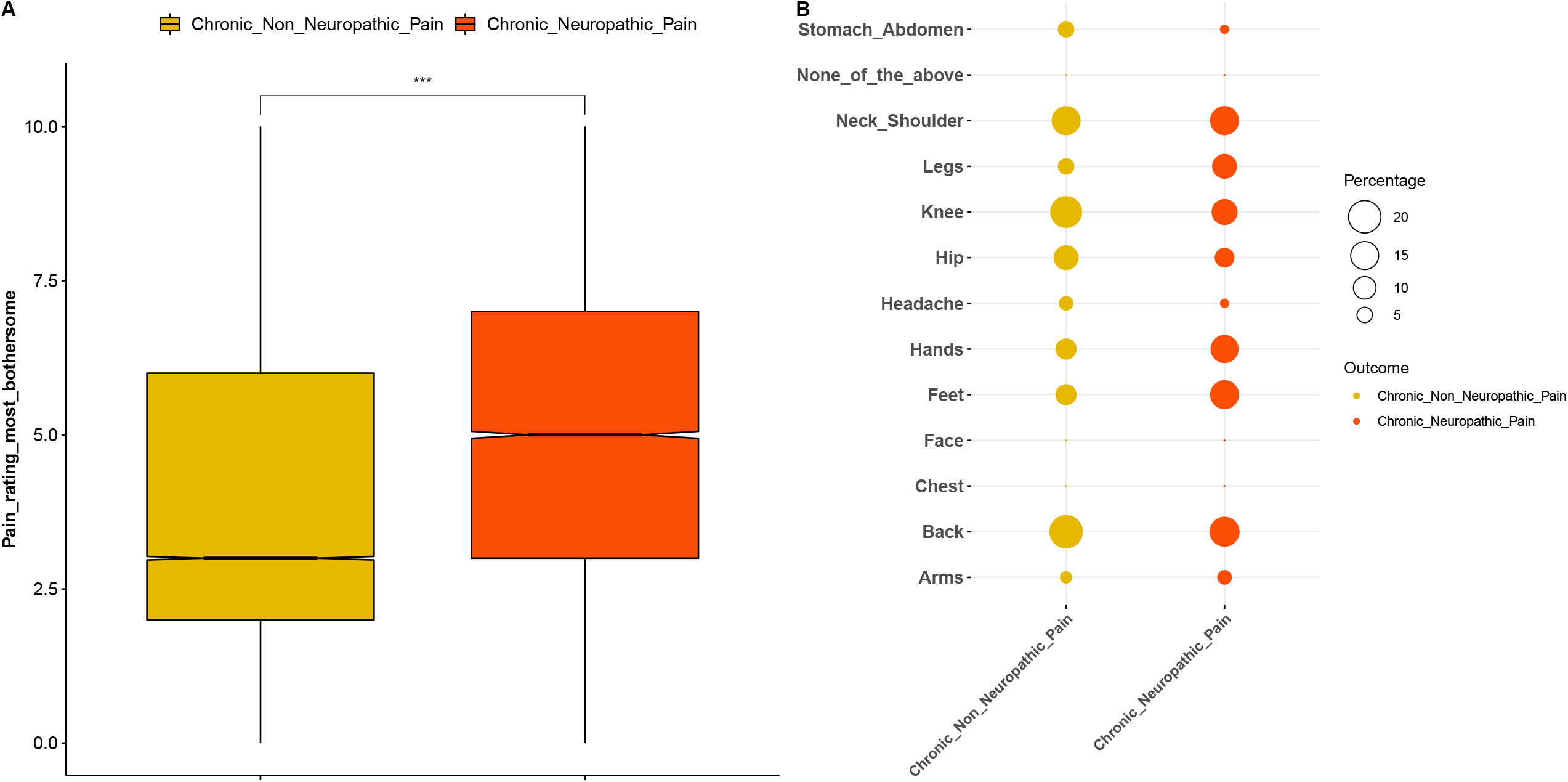
Pain rating and location of the most bothersome pain. A: Boxplots show the pain rating for the self-reported location of the most bothersome pain for both painful groups, NeuP vs Non-NeuP. B: Dotplot shows the frequencies for the self-reported location of the most bothersome pain for the two painful groups. Rates are coded in the size of the dot. P.value < 0.001 is coded as ***, p.value < 0.01 is coded as ** 0.01, p.value < 0.05 is coded as *.

**Figure 4.**
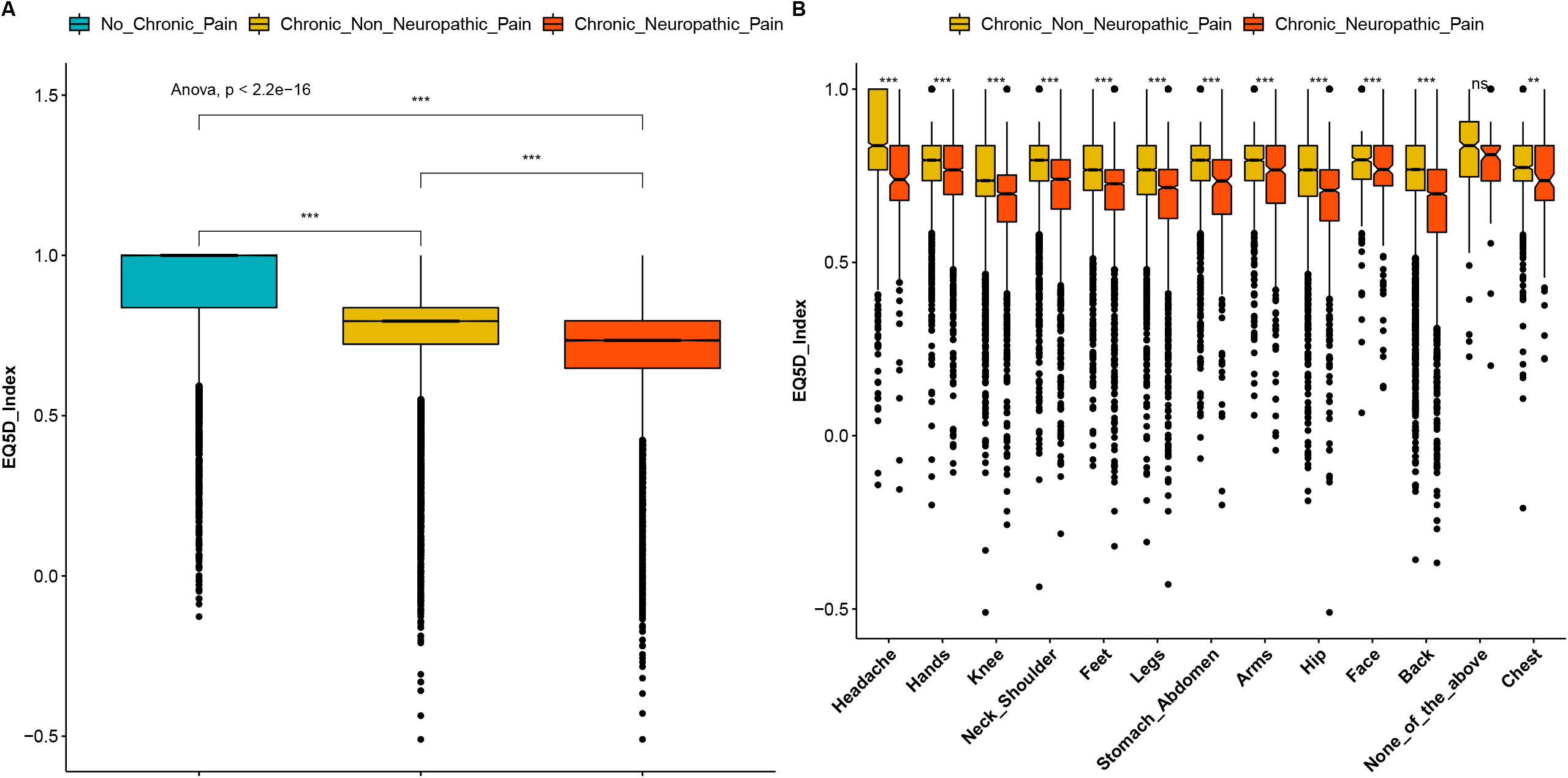
The impact of pain in quality of life. A: Boxplots show the EQ5D index across the three groups. B: Boxplots show the EQ5D for both painful groups across all locations reported as the ones having the most bothersome pain. Omnibus ANOVAs are labelled In the plot and are followed-up by t-tests between groups. P.value < 0.001 is coded as ***, p.value < 0.01 is coded as ** 0.01, p.value < 0.05 is coded as *. Post-hoc tests are shown in supplementary figure 3.

**Figure 5.**
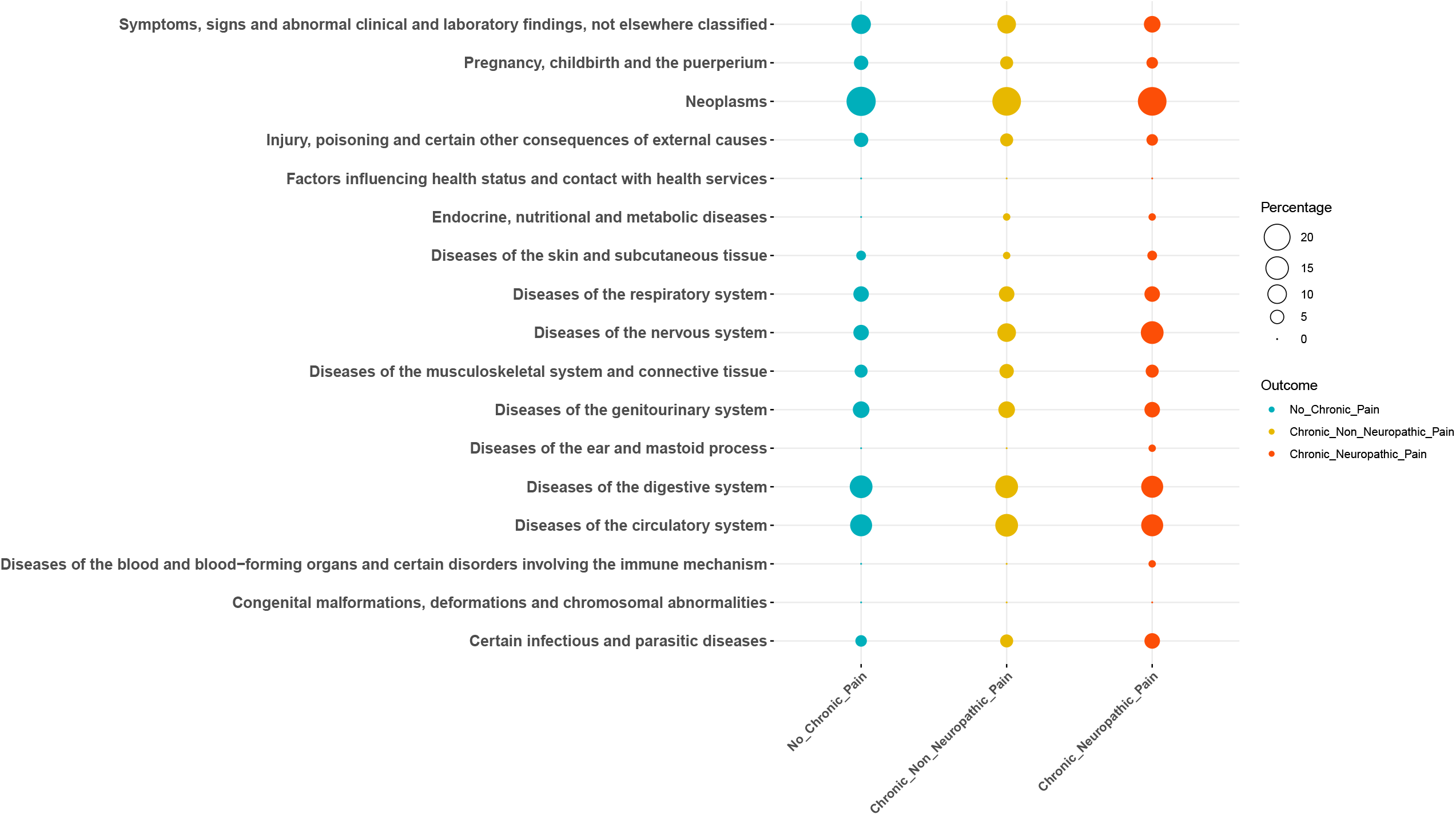
Frequencies of diseases across the three groups. The rates are coded in the size of the dot. Group allocation is colour coded. Post-hoc tests are shown in supplementary figure 4.

Focusing on people with chronic pain (both NeuP and Non-NeuP), we observed that DN4 scores were significantly associated with the pain severity rating and were higher for participants with higher pain ratings for the most bothersome pain, figure 6A (p.value < 0.001). The highest DN4 scores were those of participants with the most bothersome pain in the feet and legs, followed by hands and face, figure 6B. People with pain in both feet had significantly higher DN4 scores than people with pain only in one foot (p.value < 0.001), figure 6C. Pain severity ratings for the most bothersome location were higher in participants with higher BMI scores (figure 7A), lower quality of life (figure 7B), and/or who were unable to work due to sickness or disability (figure 7C) and slightly higher for among participants with higher deprivation scores (figure 7D).

**Figure 6.**
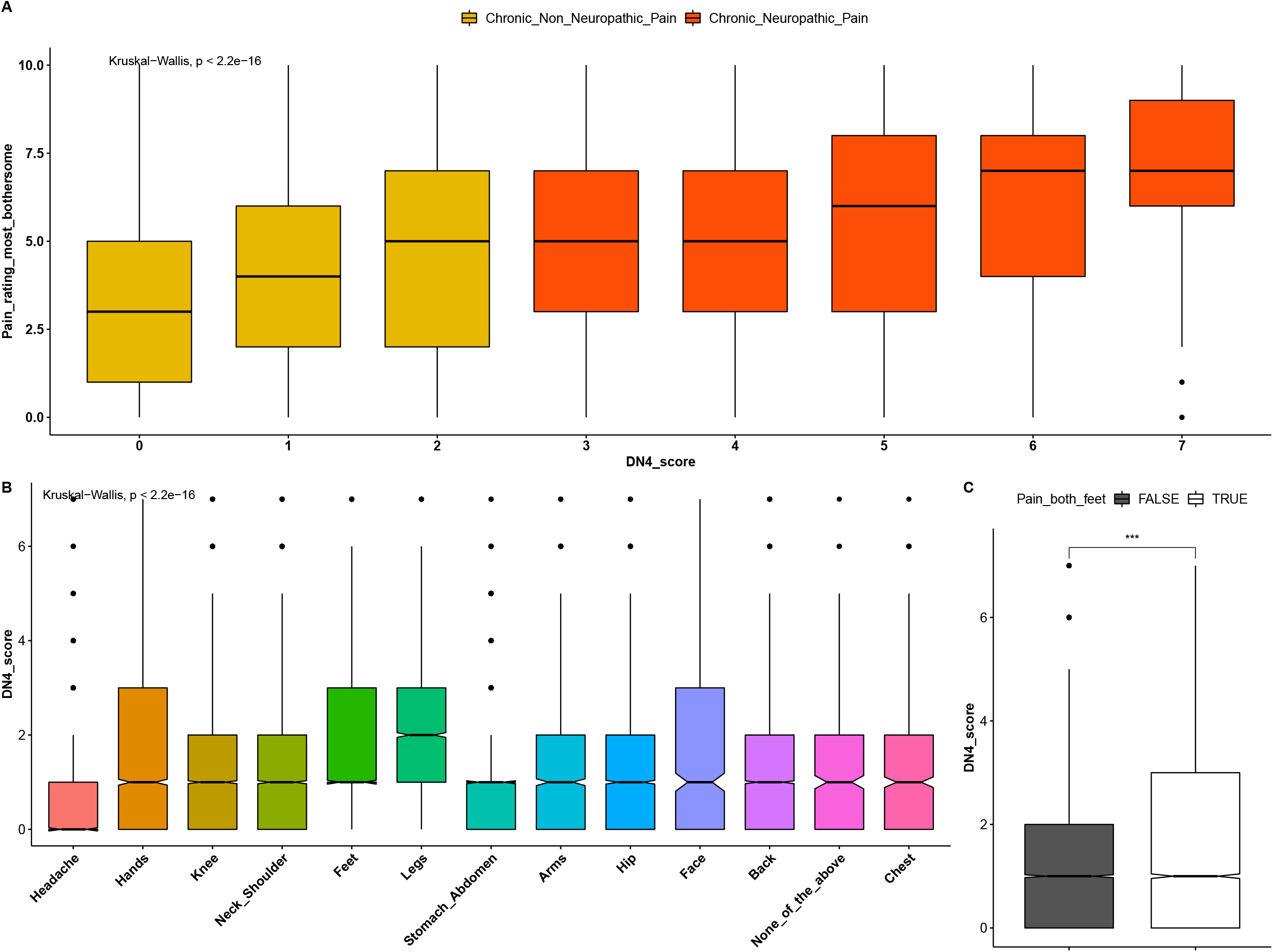
Pain rating and DN4 scores. A: Boxplots show the distribution of pain ratings for the most bothersome locations across DN4 scores for both painful groups. Group is colour coded. B: Boxplots show the DN4 score for all different locations of self-reported most bothersome pain. C: DN4 scores for people with and without pain in both feet, Omnibus Kruskall-Wallis tests are labelled in the plot. P.value < 0.001 is coded as ***, p.value < 0.01 is coded as ** 0.01, p.value < 0.05 is coded as *.

**Figure 7.**
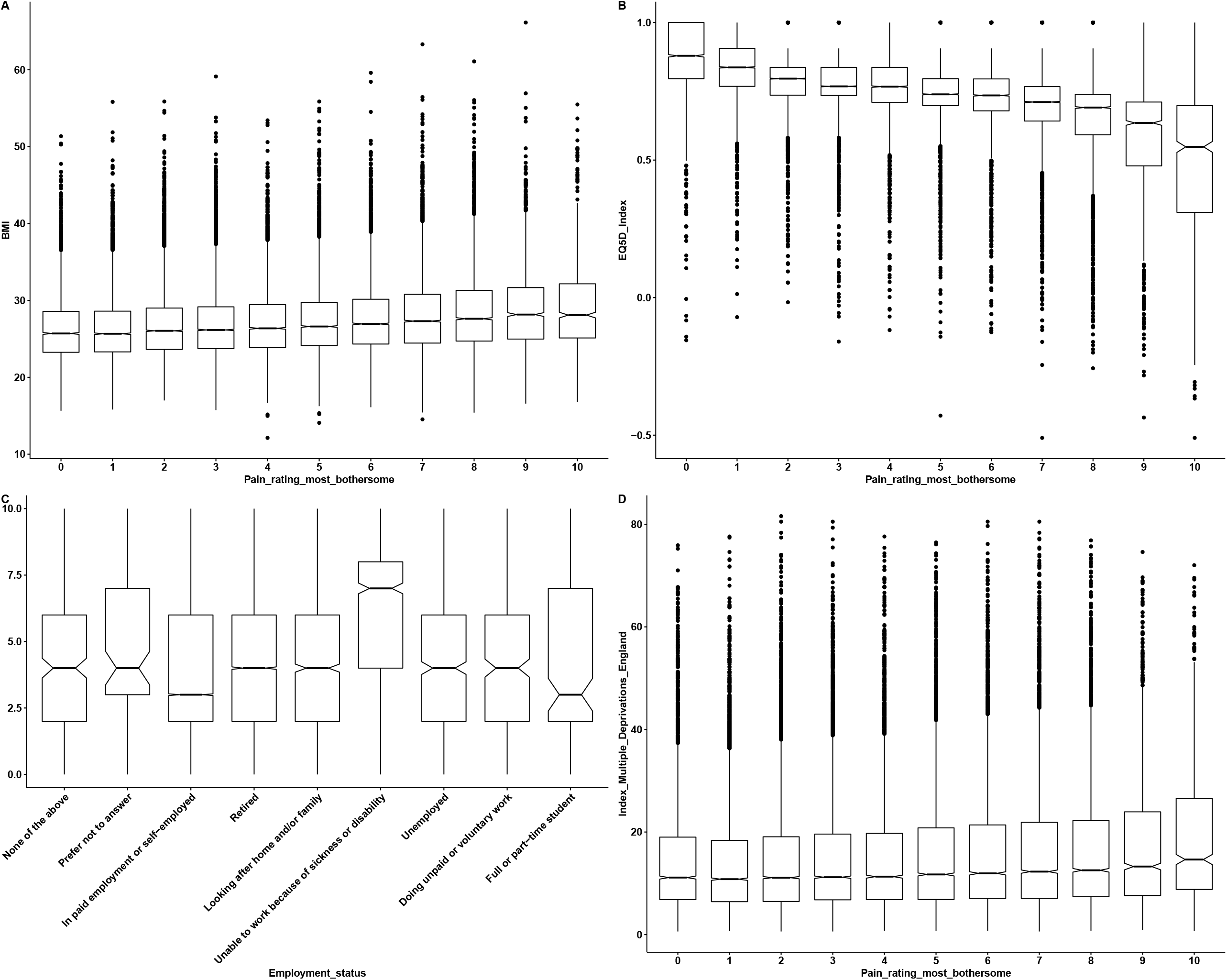
Intensity of pain versus participant characteristics. A: Boxplots show the distribution BMI for different self-reported pain intensities for the most bothersome location. B: Boxplots show the EQ5D index across self-reported pain intensities for the most bothersome locations. C: Frequencies of employment status across self-reported pain intensities for the most bothersome locations. D: Index of multiple deprivations or different self-reported pain intensities for the most bothersome location. Notched lines represent the median.

### 3.3. Multivariable modelling

In figure 8 and supplementary table 5 we present the exponentiated coefficients/odds ratios (ORs) for all terms that reached significance (one-way ANOVA p < 0.05) for the multinomial model with a three-level dependent variable considering no chronic pain as the reference condition. Both NeuP and non-neuP were strongly associated with having lower QoL (assessed using EQ5D in which a higher score reflects a higher quality of life) The presence of diabetes increased the odds ratio for NeuP and decreased them for Non-NeuP. All other self-reported conditions increased the odds ratios for both NeuP and Non-NeuP but had a larger effect for NeuP. Males had lower odds ratio for both NeuP and Non-NeuP. In addition those who held “Associate Professional and Technical Occupations” or were “Managers and Senior Officials” were more likely to report Non-NeuP. “Associate Professional and Technical Occupations”, “Process, Plant and Machine Operatives” and “Skilled Trades Occupations” increased the odds for NeuP, while “Professional Occupations” decreased the odds for NeuP.

**Figure 8.**
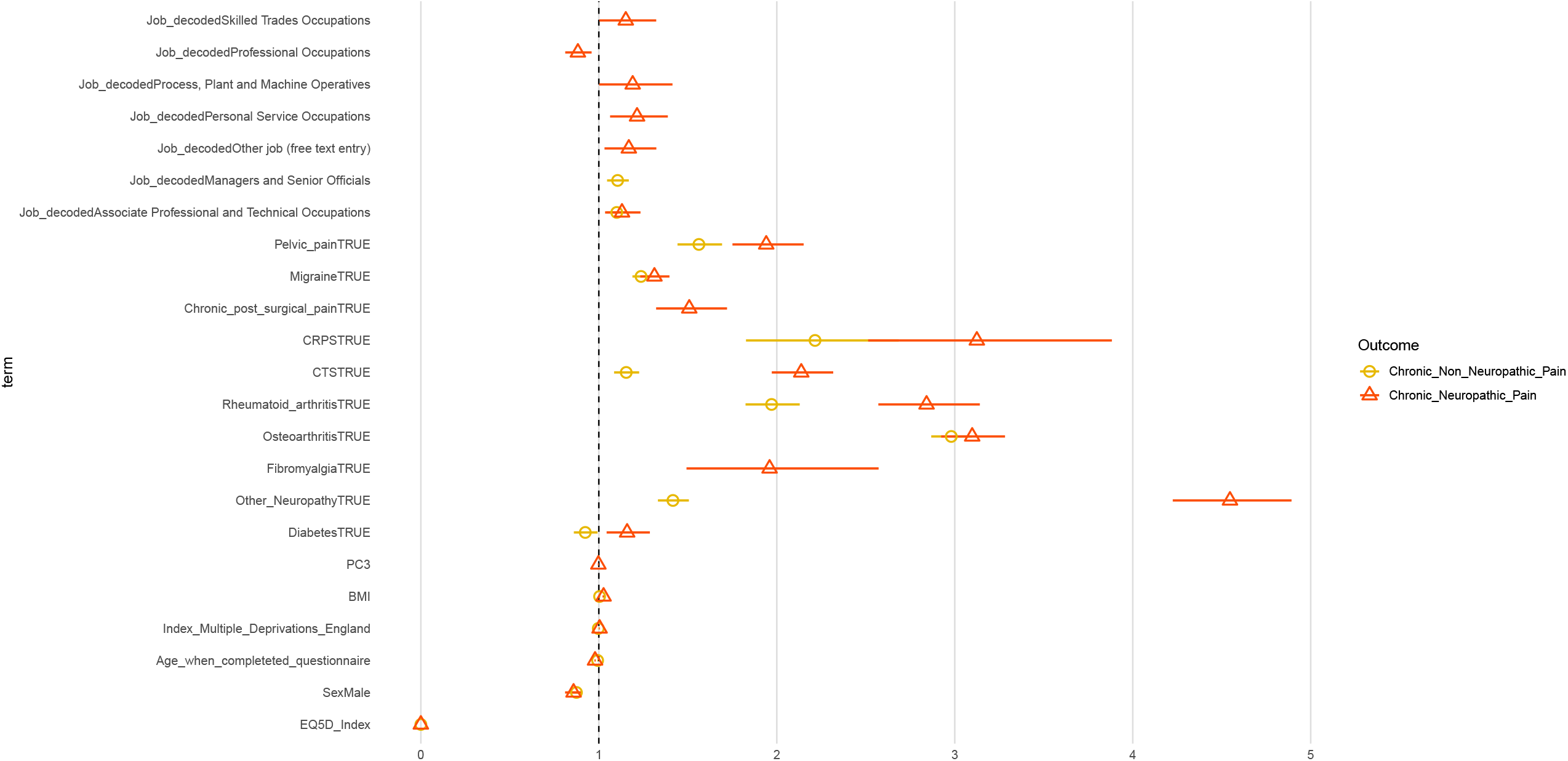
Coefficient estimates of the multinomial logit model. Dots represent the exponentiated coefficient estimates, i.e. odds ratios, and lines show the 95% confidence interval for all terms with an one-way ANOVA p.value < 0.05. Odds ratios are calculated for NeuP and Non-NeuP against the reference level NoCP.

In figure 9 and supplementary table 6 we present the ORs for a binomial model with a two-level dependent variable, NeuP versus Non-NeuP. Male participants had a higher OR for NeuP vs Non-NeuP. Higher BMI and lower age were associated with an increased OR for NeuP. Most bothersome pain in the feet was associated with an increased OR for NeuP, whereas most bothersome pain in the abdomen, shoulder, knee, hip. Back, chest and headache was associated with reduced OR for NeuP. “Personal Service Occupations” was associated with increased OR for NeuP and “Professional Occupations” with decreased OR for NeuP.

**Figure 9.**
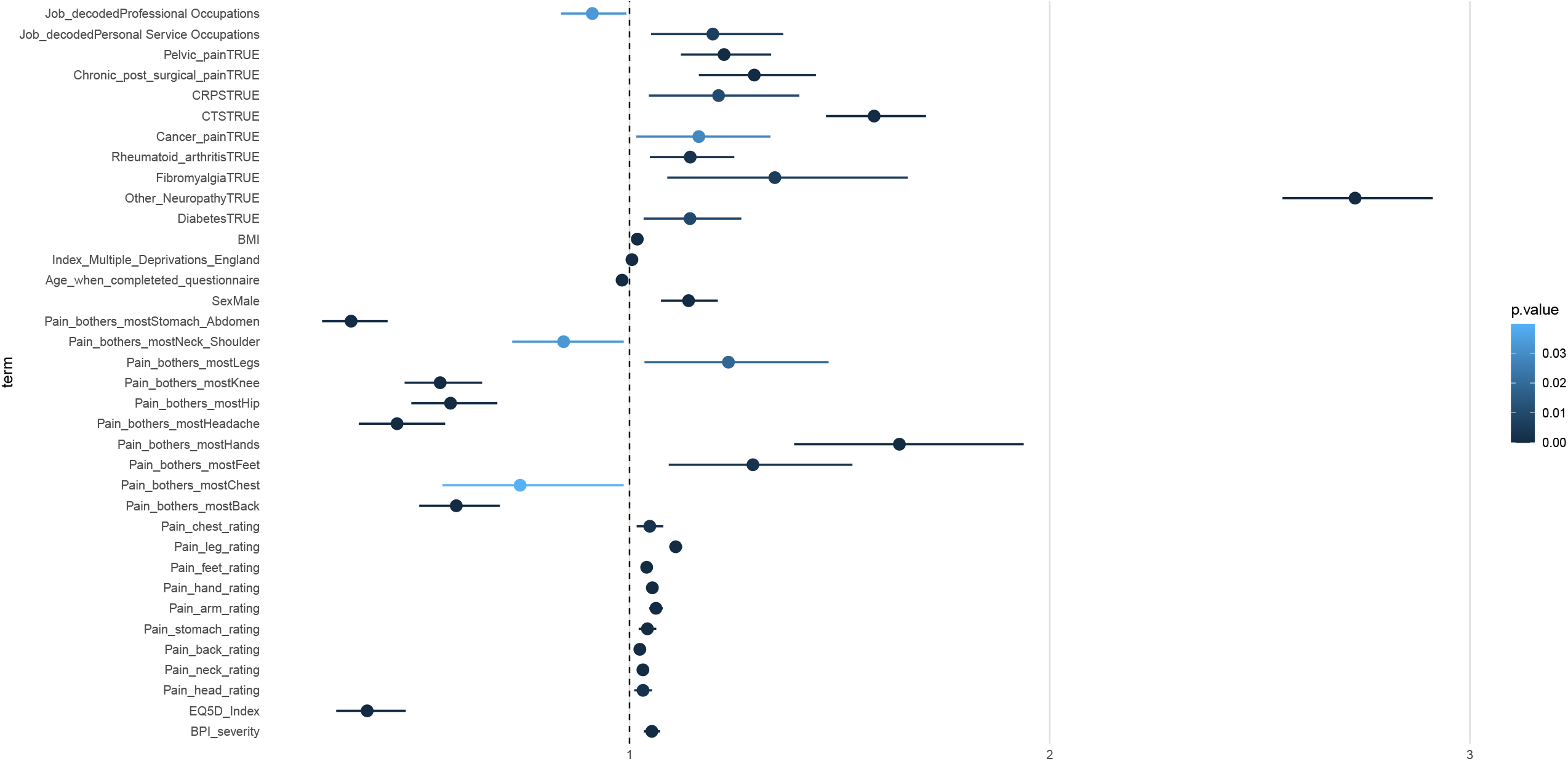
Coefficient estimates of the binomial logit model for the two painful groups. Dots represent the exponentiated coefficient estimates, i.e odds ratios, and lines show the 95% confidence interval for all terms with an one-way ANOVA p.value < 0.05. Odds ratios are calculated for NeuP against the reference level Non-NeuP (constituting the sum of Non-NeuP and NoCP). The p.value is colour coded.

## 4. Discussion

### 4.1. Summary

In this large cross-sectional study of middle-aged adults in UK Biobank, the prevalence of NeuP was 9.2%, making up 18.1% of those with chronic pain. After adjusting for confounding variables, factors associated with NeuP compared to NoCP were worse health-related quality of life, increased social deprivation, having manual or personal service type occupations, not having a professional occupation and younger age. These factors were also associated with NeuP compared to non-NeuP, as were pain in the upper and lower extremities, worse pain location intensities and higher BMI. Female sex was associated with NeuP compared to NoCP, but male sex was associated with NeuP compared to non-NeuP. NeuP co-morbidities included expected associations such as neuropathy and diabetes but also other pain (pelvic pain, migraine and post-surgical pain) and musculoskeletal disorders (rheumatoid arthritis, osteoarthritis and fibromyalgia).

### 4.2. Interpretation

The published range of prevalence estimates for NeuP in the general population is 3.2-17.9% [9,14,25,32,38,51,54,60–62]. In the UK, two studies reported a prevalence of 8.2% [50] and 9.3% [48] respectively. These two figures are similar to the current study (9.2%). It should be noted though that these studies did not cover Wales or Northern Ireland and the current study did not cover Northern Ireland. The wide range of prevalences reported is partly due to differences in case ascertainment and definition. In Japan, the relatively low prevalence of 3.2% can be explained by the stringent case definition used, which included a longer duration of pain (6 months compared to 3 months) and pain intensity of 5 or more out of 10 [32]. In studies that used the same definition as the current one, the prevalence of NeuP was lower in France (6.9%; [9]) and higher in Canada (16.1% and 17.9%; [51,54]) and Morocco (10.6%; [25]), though these studies have a wider age range than the current study, which is predominantly middle and older aged.

The current study has enabled the identification of associations with a greater level of precision than previous studies. First, people with NeuP have worse health-related quality of life than people without NeuP [3,32,49,51]. Most of the studies reporting this finding used only univariate hypothesis tests, i.e. considering only one variable at the time, which does not take potential confounding factors into account, such as differing pain intensity [38,62], in contrast to the current study.

Second, this study supports previous findings that pain intensity is more severe in people with NeuP than people with non-NeuP [9,25,32,38,48,50,51,60,62]. Part of the possible reason for this is the relative difficulty in treating NeuP. It has previously been reported that significantly fewer people with NeuP rate their prescribed treatment (medication or other forms of therapy) as “very successful” [60].

Third, people with NeuP were more likely to report pain in their hands, legs or feet [9,25,32,50,62], whilst people with non-NeuP were more likely to report pain in their stomach or abdomen, neck or shoulder, knees, hips, head, chest and back [25,50,62]. Common aetiologies of NeuP including diabetes, surgery, carpal tunnel syndrome, complex regional pain syndrome and cancer often affect the upper and lower extremities. This is supported by the association of these comorbidities independently with NeuP.

In addition to these findings, we identified some novel associations with NeuP in the general population. This includes rheumatoid arthritis. Whilst arthritis is generally considered to be non-neuropathic in nature, there is growing evidence of a neuropathic component to this cause of pain [17,33,34,45]. Furthermore, rheumatoid arthritis can predispose to NeuP disorders such as carpal tunnel syndrome [58], vasculitic neuropathy [24] and spinal cord compression [15]. However, a previous study found no association with NeuP [22]. It is also unclear in this study whether rheumatoid arthritis is directly related to the NeuP reported. As individuals can suffer from pain in multiple locations, it is possible that rheumatoid arthritis could be present in a person with NeuP in another location with a different cause.

Another novel finding of this study is that BMI was significantly higher in people with NeuP than in people with non-NeuP. Despite not being previously reported in the general population, this association has been reported in specific populations with NeuP including diabetic peripheral neuropathy [44] and rheumatoid arthritis [33]. Higher BMI is associated with increased risk of common conditions causing NeuP including diabetes [57] and cardiovascular disease [37], whilst obesity can place increased strain on the joints, exacerbating painful symptoms in rheumatoid arthritis [4].

This study has identified some associations that conflicted with the literature. Male sex was associated with increased risk of having NeuP compared to non-NeuP, whereas female sex was associated with increased of having NeuP compared to NoCP. Female sex has been consistently associated with NeuP when compared to non-NeuP in the literature, in both adjusted [9,50,60,62] and unadjusted analysis [25,48,51]. A notable exception is a Canadian study, which also used the DN4 [54]. However, a separate analysis in that study using the S-LANSS found a significant association with female sex, highlighting the heterogeneity that can arise from different phenotyping methods.

Younger age, analysed as a continuous variable, was significantly associated with NeuP. Previous studies have mostly analysed age as a categorical variable and found that NeuP is more prevalent in older people [9,25,32,50,54], though some have reported no association [48,51,60]. A study conducted in France identified a peak prevalence of NeuP in the 50-64 year group, with the under 25, 25-34 and 65-74 year groups all having lower prevalences in comparison [9]. As the age range of the cohort for the current study was 49-83 (Q1 = 61, Q3 = 73) years (taking into account the time between initial recruitment for UK Biobank and completion of the chronic pain phenotyping survey), the age group expected to have the higher prevalence of NeuP (the 50-64 year group) were amongst the younger participants of UK Biobank. This could explain the findings in this study.

Being in a personal service, skilled trade or plant and machine operative job and not being in a professional occupation, were associated with NeuP. A previous study in France found that NeuP was significantly more prevalent in manual workers, farmers, retirees and other non-working people, compared to those in managerial positions [9]. These are occupations in which people are likely to require repetitive use of hands and feet, which could make nerve damage more likely. However, it should be noted that a study in Hong Kong found no association of NeuP with manual, clerical or service industry jobs when compared to professional jobs [60].

### 4.3. Strengths and Limitations

The main strength of this study is the NeuP sample size (n=13,747) which is more than 5 times that of the largest previous study in the general population [14]. This was achieved by way of a pain phenotyping survey that used validated questionnaires, such as the DN4, BPI, CPG and MNSI. This will allow direct comparison with other studies of NeuP, notably DOLORisk [27,40] and the Pain in Neuropathy Study (PiNS) [47]. Additionally, we conducted multivariate regression analysis on a wide range of factors relating to pain, which enabled us to control for confounding factors. This compares to previous studies of NeuP in the general population where regression has not been conducted [14,25,32,48,51,61]. The response rate of approximately 50% compares favourably with previous studies [9,14,61,62,25,32,38,48,50,51,54,60].

In contrast, this study also has some limitations. Our definition of NeuP relies on a self-completed screening tool for NeuP, which does not meet the grading system for “probable” or “definite” NeuP [19]. This includes clinical examination which is clearly not feasible in a population survey. Despite this, there are a number of previous studies that have used a similar definition to the current one and can be used as direct comparisons [9,25,51,54].

The study was cross-sectional, which means it is not possible to determine whether the relationships between NeuP and the independent variables reported are causal [31]. However, for some of these variables, particularly the non-modifiable ones such as age and sex, the temporal relationship can be inferred. There are currently plans for UK Biobank to repeat the chronic pain survey in 2023 and this will present an opportunity to conduct further longitudinal analysis to identify incidence and predictors of NeuP onset.

A further weakness of the study is that the UK Biobank is limited to white middle-aged adults which means that the cohort is not completely representative of the general population. It has also been suggested that the UK Biobank under-represents people from more socioeconomically deprived areas, as well as people who are obese, smoke, drink alcohol and self-report certain health conditions [21]. Since NeuP has previously been associated with these health-related states, our estimate of the prevalence of NeuP may be conservative [10,33,39,44]. We also found an over-representation of females, younger age, lower BMI and less social deprivation in those participating in the pain phenotyping survey compared to those who did not participate, suggesting an element of ascertainment bias. However, it should be noted that whilst these observations reached significance statistically, this may reflect the fact that the study was highly powered to detect even small differences between groups.

Unlike in a number of previous studies [38,60,62], we were unable to analyse the treatments participants had received for their pain, either through healthcare professionals or through self-medication. Therefore, this should be a focus of future work. Similarly, future studies will explore the wealth of genetic data that is available within UK Biobank, both to validate recent findings and to potential identify novel associations [55,56].

### 4.4. Conclusions

This study represents the largest epidemiological study of NeuP to date and confirms that the disorder is common in the general population and places a significantly greater burden on people compared to those without the disorder (whether measuring those with chronic non-neuropathic pain, or those reporting no chronic pain). In addition, it has identified sections of the population with particular demographic and clinical characteristics that are at increased risk of having chronic NeuP, some of which may be amenable to targeted prevention. These findings will be of particular interest to healthcare professionals who will be able to influence clinical policy for preventing and treating the disorder.

## Data Availability

Data for this study was obtained from the UK biobank for project Risk factors for chronic pain, Application ID: 49572.

https://www.ukbiobank.ac.uk/

## Acknowledgements

This work was supported by Diabetes UK under grant agreement 19/0005984, the European Union’s Horizon 2020 research and innovation programme under grant agreement No 633491 (DOLORisk) and the MRC and Versus Arthritis funding to the PAINSTORM consortium as part of the Advanced Pain Discovery Platform (MR/W002388/1). No funding body had any role in the design of the study and collection, analysis, and interpretation of data and in writing the manuscript. A.C.T is supported by Academy of Medical Sciences Starter Grant SGL022\1086, and is a Honorary Senior Research Fellow and Carnegie-Wits Diaspora Fellow at the Brain Function Research Group, School of Physiology, Faculty of Health Sciences, University of the Witwatersrand, Johannesburg. D.L.B and G.B are supported by the Wellcome Trust, grant reference 223149/Z/21/Z. For the purpose of open access, the authors have applied a CC BY public copyright licence to any Author Accepted Manuscript version arising from this submission.

## Ethics

Data for this study was obtained from the UK biobank for project “Risk factors for chronic pain”, Application ID: 49572. UK Biobank has approval from the North West Multi-centre Research Ethics Committee (MREC) as a Research Tissue Bank (RTB) approval, REC reference: 21/NW/0157, IRAS project ID: 299116. This approval means that researchers do not require separate ethical clearance and can operate under the RTB approval.

