## Supplemental material for "The epidemiology of neuropathic pain: an analysis of prevalence and associated factors in UK Biobank"

| | Item No | Recommendation | Page No |
|---|---|---|---|
| **Title and abstract** | 1 | (*a*) Indicate the study's design with a commonly used term in the title or the abstract | 1 |
| | | (*b*) Provide in the abstract an informative and balanced summary of what was done and what was found | 2 |
| **Introduction** | | | |
| Background/rationale | 2 | Explain the scientific background and rationale for the investigation being reported | 3 |
| Objectives | 3 | State specific objectives, including any prespecified hypotheses | 3 |
| **Methods** | | | |
| Study design | 4 | Present key elements of study design early in the paper | 4 |
| Setting | 5 | Describe the setting, locations, and relevant dates, including periods of recruitment, exposure, follow-up, and data collection | 4 |
| Participants | 6 | (*a*) Give the eligibility criteria, and the sources and methods of selection of participants | 4 |
| Variables | 7 | Clearly define all outcomes, exposures, predictors, potential confounders, and effect modifiers. Give diagnostic criteria, if applicable | 4-5 |
| Data sources/ measurement | 8* | For each variable of interest, give sources of data and details of methods of assessment (measurement). Describe comparability of assessment methods if there is more than one group | 4-5 |
| Bias | 9 | Describe any efforts to address potential sources of bias | NA |
| Study size | 10 | Explain how the study size was arrived at | 5 |
| Quantitative variables | 11 | Explain how quantitative variables were handled in the analyses. If applicable, describe which groupings were chosen and why | 5 |
| Statistical methods | 12 | (*a*) Describe all statistical methods, including those used to control for confounding | 5-6 |
| | | (*b*) Describe any methods used to examine subgroups and interactions | NA |
| | | (*c*) Explain how missing data were addressed | 6 |
| | | (*d*) If applicable, describe analytical methods taking account of sampling strategy | NA |
| | | (*e*) Describe any sensitivity analyses | NA |
| **Results** | | | |
| Participants | 13* | (a) Report numbers of individuals at each stage of study—eg numbers potentially eligible, examined for eligibility, confirmed eligible, included in the study, completing follow-up, and analysed | 7 |
| | | (b) Give reasons for non-participation at each stage | NA |
| | | (c) Consider use of a flow diagram | NA |
| Descriptive data | 14* | (a) Give characteristics of study participants (eg demographic, clinical, social) and information on exposures and potential confounders | 7 |
| | | (b) Indicate number of participants with missing data for each variable of interest | Table S2 |
| Outcome data | 15* | Report numbers of outcome events or summary measures | 7 |

| | | | |
|---|---|---|---|
| | | | Tables S3-S4 |
| Main results | 16 | (*a*) Give unadjusted estimates and, if applicable, confounder-adjusted estimates and their precision (eg, 95% confidence interval). Make clear which confounders were adjusted for and why they were included | 7-8<br>Figures 1-9<br>Table S5<br>Figure S3-S6 |
| | | (*b*) Report category boundaries when continuous variables were categorized | NA |
| | | (*c*) If relevant, consider translating estimates of relative risk into absolute risk for a meaningful time period | NA |
| Other analyses | 17 | Report other analyses done—eg analyses of subgroups and interactions, and sensitivity analyses | NA |
| **Discussion** | | | |
| Key results | 18 | Summarise key results with reference to study objectives | 9 |
| Limitations | 19 | Discuss limitations of the study, taking into account sources of potential bias or imprecision. Discuss both direction and magnitude of any potential bias | 11 |
| Interpretation | 20 | Give a cautious overall interpretation of results considering objectives, limitations, multiplicity of analyses, results from similar studies, and other relevant evidence | 9-10 |
| Generalisability | 21 | Discuss the generalisability (external validity) of the study results | 9-11 |
| **Other information** | | | |
| Funding | 22 | Give the source of funding and the role of the funders for the present study and, if applicable, for the original study on which the present article is based | 12 |

*Give information separately for exposed and unexposed groups.

Supplementary table 1: STROBE Statement—Checklist of items that should be included in reports of ***cross-sectional studies***

**Note:** An Explanation and Elaboration article discusses each checklist item and gives methodological background and published examples of transparent reporting. The STROBE checklist is best used in conjunction with this article (freely available on the Web sites of PLoS Medicine at http://www.plosmedicine.org/, Annals of Internal Medicine at http://www.annals.org/, and Epidemiology at http://www.epidem.com/). Information on the STROBE Initiative is available at www.strobe-statement.org.

| Variable | Missing values percentages % |
|---|---|
| BPI_severity | 0.7 |
| BPI_interference | 0.8 |
| EQ5D_Index | 0 |
| EQ5D_Health_today | 0.2 |
| Pain_both_feet | 65 |
| Pain_head_rating | 0.2 |
| Pain_face_rating | 0.2 |
| Pain_neck_rating | 0.2 |
| Pain_back_rating | 0.2 |
| Pain_stomach_rating | 0.3 |
| Pain_hip_rating | 0.4 |
| Pain_knee_rating | 0.2 |
| Pain_arm_rating | 0.2 |
| Pain_hand_rating | 0.2 |
| Pain_feet_rating | 0.2 |
| Pain_leg_rating | 0.3 |
| Pain_chest_rating | 0.3 |
| Pain_bothers_most | 17.4 |
| Pain_rating_most_bothersome | 18.2 |
| Sex | 0 |
| Ethnic_background | 0 |
| Age_when_completeted_questionnaire | 0 |
| Index_Multiple_Deprivations_England | 12.9 |
| Index_Multiple_Deprivations_Wales | 96.2 |
| Index_Multiple_Deprivations_Scotland | 93.5 |
| BMI | 0.3 |
| Employment_status | 0 |
| PC1 | 2 |
| PC2 | 2 |
| PC3 | 2 |
| Diabetes | 0.3 |
| Other_Neuropathy | 2.6 |
| Fibromyalgia | 2.6 |
| CFS_ME | 0 |
| Osteoarthritis | 3.9 |
| Rheumatoid_arthritis | 5.2 |
| Cancer_pain | 0.7 |
| CTS | 2.4 |

| | |
|---|---|
| CRPS | 6.4 |
| Chronic_post_surgical_pain | 1 |
| Gout | 0.4 |
| Migraine | 0.5 |
| Pelvic_pain | 1.1 |
| Job_decoded | 27.4 |
| ICD_decoded_toplevel | 61.3 |
| Ethnic_background_grouped | 1 |

Supplementary table 2: Percentages of missing values for all variables considered in both the descriptive hypothesis testing and multivariable modelling.

| Pain_experience | | Completed | Not_Completed | Total | p |
|---|---|---|---|---|---|
| Sex | Female | 95002 (56.8) | 178326 (53.2) | 273328 (54.4) | <0.001 |
| | Male | 72201 (43.2) | 156884 (46.8) | 229085 (45.6) | |
| Age_at_recruitment | Mean (SD) | 55.7 (7.7) | 56.9 (8.3) | 56.5 (8.1) | <0.001 |
| BMI | Mean (SD) | 26.8 (4.6) | 27.8 (4.9) | 27.4 (4.8) | <0.001 |
| Index_Multiple_Deprivations_England | Mean (SD) | 15.2 (12.0) | 18.9 (14.8) | 17.7 (14.0) | <0.001 |

Supplementary table 3: Demographics, BMI and relative deprivation for the group that has completed versus not completed the pain re-phenotyping in UKB.

| **DN4_completed** | | **Completed** | **Not_Completed** | **Total** |
|---|---|---|---|---|
| Sex | Female | 44755 (58.8) | 50247 (55.2) | 95002 (56.8) |
| | Male | 31340 (41.2) | 40861 (44.8) | 72201 (43.2) |
| Age_at_recruitment | Mean (SD) | 55.9 (7.6) | 55.6 (7.7) | 55.7 (7.7) |
| BMI | Mean (SD) | 27.1 (4.7) | 26.6 (4.5) | 26.8 (4.6) |
| Index_Multiple_Deprivations_ England | Mean (SD) | 15.2 (12.0) | 15.2 (12.1) | 15.2 (12.0) |

Supplementary table 4: Demographics, BMI and relative deprivation for the group that has completed pain re-phenotyping and has a fully completed DN4 versus an incomplete DN4 questionnaire.

| Outcome level | Coefficient | Odds-Ratio | std.error | statistic | df | p.value | 95%CI.low | 95%CI.high |
|---|---|---|---|---|---|---|---|---|
| Non-NeuP | (Intercept) | 2814.8467 | 0.1776 | 44.7135 | 941.5233 | <0.0001 | 1986.3568 | 3988.8916 |
| Non-NeuP | EQ5D_Index | 1.00E-04 | 0.0725 | -130.7751 | 105520.5909 | <0.0001 | 1.00E-04 | 1.00E-04 |
| Non-NeuP | SexMale | 0.8741 | 0.0171 | -7.8598 | 103496.7438 | <0.0001 | 0.8453 | 0.9039 |
| Non-NeuP | Age_when_completeted_questionnaire | 0.9932 | 0.0011 | -6.1637 | 100056.8677 | <0.0001 | 0.9911 | 0.9954 |
| Non-NeuP | Index_Multiple_Deprivations_England | 0.9969 | 7.00E-04 | -4.2404 | 1176.757 | <0.0001 | 0.9955 | 0.9984 |
| Non-NeuP | BMI | 1.0044 | 0.0019 | 2.2899 | 98869.0434 | 0.022 | 1.0006 | 1.0081 |
| Non-NeuP | DiabetesYes | 0.9238 | 0.0367 | -2.1611 | 69964.2693 | 0.0307 | 0.8598 | 0.9927 |
| Non-NeuP | Other_NeuropathyYes | 1.4164 | 0.0312 | 11.1409 | 54693.5849 | <0.0001 | 1.3322 | 1.5058 |
| Non-NeuP | OsteoarthritisYes | 2.9804 | 0.0196 | 55.5868 | 14365.2578 | <0.0001 | 2.8678 | 3.0974 |
| Non-NeuP | Rheumatoid_arthritisYes | 1.9707 | 0.0393 | 17.2457 | 9026.8154 | <0.0001 | 1.8244 | 2.1286 |
| Non-NeuP | CTSYes | 1.154 | 0.0311 | 4.6071 | 25941.8873 | <0.0001 | 1.0858 | 1.2264 |
| Non-NeuP | CRPSYes | 2.2147 | 0.0982 | 8.0986 | 7613.2815 | <0.0001 | 1.827 | 2.6848 |
| Non-NeuP | MigraineYes | 1.2372 | 0.0203 | 10.4721 | 74672.0766 | <0.0001 | 1.1889 | 1.2875 |
| Non-NeuP | Pelvic_painYes | 1.5624 | 0.0408 | 10.9325 | 50868.6053 | <0.0001 | 1.4422 | 1.6925 |
| Non-NeuP | Job_decodedAssociate Professional and Technical Occupations | 1.1006 | 0.0274 | 3.5057 | 107514.9586 | 5.00E-04 | 1.0432 | 1.1613 |
| Non-NeuP | Job_decodedManagers and Senior Officials | 1.1056 | 0.0281 | 3.5731 | 106232.821 | 4.00E-04 | 1.0464 | 1.1682 |
| NeuP | (Intercept) | 3693.1546 | 0.2664 | 30.8299 | 2994.4386 | <0.0001 | 2190.352 | 6227.0312 |
| NeuP | EQ5D_Index | <0.0001 | 0.1029 | -114.0615 | 100771.4724 | <0.0001 | 0 | 0 |
| NeuP | SexMale | 0.858 | 0.0291 | -5.2579 | 98512.6293 | <0.0001 | 0.8105 | 0.9084 |
| NeuP | Age_when_completeted_questionnaire | 0.9788 | 0.0019 | -11.5564 | 93625.0376 | <0.0001 | 0.9753 | 0.9824 |
| NeuP | Index_Multiple_Deprivations_England | 1.0043 | 0.0011 | 3.8351 | 1300.2144 | 1.00E-04 | 1.0021 | 1.0065 |
| NeuP | BMI | 1.0266 | 0.0029 | 9.2005 | 87282.312 | 0 | 1.0209 | 1.0324 |
| NeuP | PC3 | 0.9974 | 0.0013 | -2.0472 | 1672.1068 | 0.0408 | 0.9949 | 0.9999 |
| NeuP | DiabetesYes | 1.1591 | 0.0533 | 2.7669 | 62976.3419 | 0.0057 | 1.044 | 1.2868 |
| NeuP | Other_NeuropathyYes | 4.5463 | 0.0374 | 40.4863 | 34719.3456 | <0.0001 | 4.2249 | 4.8921 |
| NeuP | FibromyalgiaYes | 1.9595 | 0.1388 | 4.8478 | 6243.2144 | <0.0001 | 1.4928 | 2.572 |
| NeuP | OsteoarthritisYes | 3.0978 | 0.0295 | 38.3093 | 12973.2608 | <0.0001 | 2.9237 | 3.2823 |
| NeuP | Rheumatoid_arthritisYes | 2.8414 | 0.0511 | 20.4439 | 8466.822 | <0.0001 | 2.5707 | 3.1407 |
| NeuP | CTSYes | 2.1374 | 0.0411 | 18.4717 | 8976.3611 | <0.0001 | 1.9719 | 2.3169 |
| NeuP | CRPSYes | 3.1239 | 0.1109 | 10.2717 | 3008.3319 | <0.0001 | 2.5134 | 3.8827 |
| NeuP | Chronic_post_surgical_painYes | 1.5081 | 0.0672 | 6.1154 | 25026.9388 | <0.0001 | 1.3221 | 1.7204 |
| NeuP | MigraineYes | 1.3124 | 0.0318 | 8.5515 | 75644.3979 | <0.0001 | 1.2331 | 1.3968 |
| NeuP | Pelvic_painYes | 1.9406 | 0.0525 | 12.6392 | 50596.0959 | <0.0001 | 1.751 | 2.1507 |
| NeuP | Job_decodedAssociate Professional and Technical Occupations | 1.1302 | 0.0448 | 2.7299 | 103102.6599 | 0.0063 | 1.0351 | 1.234 |
| NeuP | Job_decodedOther job (free text entry) | 1.1685 | 0.0634 | 2.4582 | 105447.1891 | 0.014 | 1.0321 | 1.323 |
| NeuP | Job_decodedPersonal Service Occupations | 1.2148 | 0.0679 | 2.8656 | 102021.6271 | 0.0042 | 1.0634 | 1.3878 |
| NeuP | Job_decodedProcess, Plant and Machine Operatives | 1.1903 | 0.0878 | 1.9838 | 98729.7843 | 0.0473 | 1.0021 | 1.4138 |
| NeuP | Job_decodedProfessional Occupations | 0.8823 | 0.0427 | -2.9306 | 98581.298 | 0.0034 | 0.8115 | 0.9594 |
| NeuP | Job_decodedSkilled Trades Occupations | 1.1515 | 0.0706 | 1.9974 | 102830.383 | 0.0458 | 1.0026 | 1.3224 |

Supplementary Table 5: Multinomial regression table, showing the Odds-ratio (i.e. exponentiated coefficient estimate), standard error of the estimate, F-statistic, p.value, lower and upper point of the 95% confidence interval for each coefficient and level of the outcome compared to NoCP.

| Coefficient | Odds-ratio | std.error | statistic | df | p.value | 95%CI. low | 95%CI. high |
|---|---|---|---|---|---|---|---|
| (Intercept) | 0.5102 | 0.2873 | -2.3416 | 1670.3511 | 0.0193 | 0.2904 | 0.8965 |
| BPI_severity | 1.0531 | 0.0094 | 5.515 | 25616.6125 | <0.0001 | 1.0339 | 1.0726 |
| EQ5D_Index | 0.3757 | 0.1115 | -8.7822 | 40523.5925 | <0.0001 | 0.3019 | 0.4674 |
| Pain_head_rating | 1.0321 | 0.0105 | 3.0194 | 35994.279 | 0.0025 | 1.0112 | 1.0535 |
| Pain_neck_rating | 1.0319 | 0.0072 | 4.3822 | 43648.5639 | <0.0001 | 1.0175 | 1.0465 |
| Pain_back_rating | 1.0245 | 0.0068 | 3.5842 | 34253.0164 | 3e-04 | 1.011 | 1.0382 |
| Pain_stomach_rating | 1.0426 | 0.0102 | 4.0752 | 43433.2413 | <0.0001 | 1.0219 | 1.0637 |
| Pain_arm_rating | 1.0629 | 0.0078 | 7.7764 | 39565.9359 | <0.0001 | 1.0467 | 1.0793 |
| Pain_hand_rating | 1.0543 | 0.0068 | 7.8247 | 38910.8099 | <0.0001 | 1.0404 | 1.0684 |
| Pain_feet_rating | 1.0408 | 0.0066 | 6.0281 | 38292.5995 | <0.0001 | 1.0274 | 1.0544 |
| Pain_leg_rating | 1.1099 | 0.007 | 14.9353 | 41074.079 | <0.0001 | 1.0948 | 1.1252 |
| Pain_chest_rating | 1.0482 | 0.0154 | 3.0536 | 39214.9791 | 0.0023 | 1.017 | 1.0803 |
| Pain_bothers_mostBack | 0.5877 | 0.0829 | -6.41 | 44144.5543 | <0.0001 | 0.4995 | 0.6914 |
| Pain_bothers_mostChest | 0.7397 | 0.1467 | -2.056 | 43434.0549 | 0.0398 | 0.5548 | 0.986 |
| Pain_bothers_mostFeet | 1.2937 | 0.0858 | 3.0017 | 43991.5507 | 0.0027 | 1.0935 | 1.5305 |
| Pain_bothers_mostHands | 1.6423 | 0.0845 | 5.8715 | 44439.3329 | <0.0001 | 1.3917 | 1.9381 |
| Pain_bothers_mostHeadache | 0.4467 | 0.1164 | -6.9237 | 44203.7701 | <0.0001 | 0.3556 | 0.5612 |
| Pain_bothers_mostHip | 0.574 | 0.0905 | -6.1334 | 43223.3115 | <0.0001 | 0.4807 | 0.6854 |
| Pain_bothers_mostKnee | 0.5494 | 0.0852 | -7.0269 | 44396.6958 | <0.0001 | 0.4648 | 0.6493 |
| Pain_bothers_mostLegs | 1.2354 | 0.09 | 2.3488 | 43983.1425 | 0.0188 | 1.0356 | 1.4738 |
| Pain_bothers_mostNeck_Shoulder | 0.8431 | 0.08 | -2.1335 | 44208.5026 | 0.0329 | 0.7208 | 0.9862 |
| Pain_bothers_mostStomach_Abdomen | 0.3374 | 0.1168 | -9.299 | 43461.1107 | <0.0001 | 0.2683 | 0.4242 |
| SexMale | 1.1405 | 0.0302 | 4.353 | 43941.609 | <0.0001 | 1.075 | 1.2101 |
| Age_when_completeted_questionnaire | 0.9822 | 0.0021 | -8.5646 | 40239.005 | <0.0001 | 0.9782 | 0.9863 |
| Index_Multiple_Deprivations_England | 1.0058 | 0.0011 | 5.203 | 2195.2878 | <0.0001 | 1.0036 | 1.0079 |
| BMI | 1.0185 | 0.0028 | 6.4725 | 39298.6866 | <0.0001 | 1.0129 | 1.0242 |
| DiabetesYes | 1.1439 | 0.0518 | 2.5962 | 37805.9533 | 0.0094 | 1.0335 | 1.2661 |
| Other_NeuropathyYes | 2.7268 | 0.0334 | 29.9946 | 3948.4265 | <0.0001 | 2.5537 | 2.9116 |
| FibromyalgiaYes | 1.3459 | 0.1076 | 2.7598 | 4060.3957 | 0.0058 | 1.0898 | 1.6621 |
| Rheumatoid_arthritisYes | 1.1445 | 0.0447 | 3.0193 | 3253.6347 | 0.0026 | 1.0484 | 1.2493 |
| Cancer_painYes | 1.1649 | 0.0697 | 2.1889 | 18764.395 | 0.0286 | 1.0161 | 1.3355 |
| CTSYes | 1.5821 | 0.0383 | 11.9724 | 3884.971 | <0.0001 | 1.4676 | 1.7055 |
| CRPSYes | 1.2119 | 0.075 | 2.5644 | 662.7275 | 0.0106 | 1.0461 | 1.4041 |
| Chronic_post_surgical_painYes | 1.2969 | 0.0546 | 4.7569 | 16342.2931 | <0.0001 | 1.1651 | 1.4435 |
| Pelvic_painYes | 1.2249 | 0.0447 | 4.5417 | 33827.7733 | <0.0001 | 1.1222 | 1.337 |
| Job_decodedPersonal Service Occupations | 1.1983 | 0.0668 | 2.7082 | 44034.8416 | 0.0068 | 1.0512 | 1.3659 |
| Job_decodedProfessional Occupations | 0.9115 | 0.0435 | -2.1312 | 43641.6093 | 0.0331 | 0.837 | 0.9926 |

Supplementary Table 6: Binomial regression table, showing the Odds-ratio (i.e. exponentiated coefficient estimate), standard error of the estimate, F-statistic, p.value, lower and upper point of the 95% confidence interval for each coefficient and level of the outcome compared to NoNeuP.

A

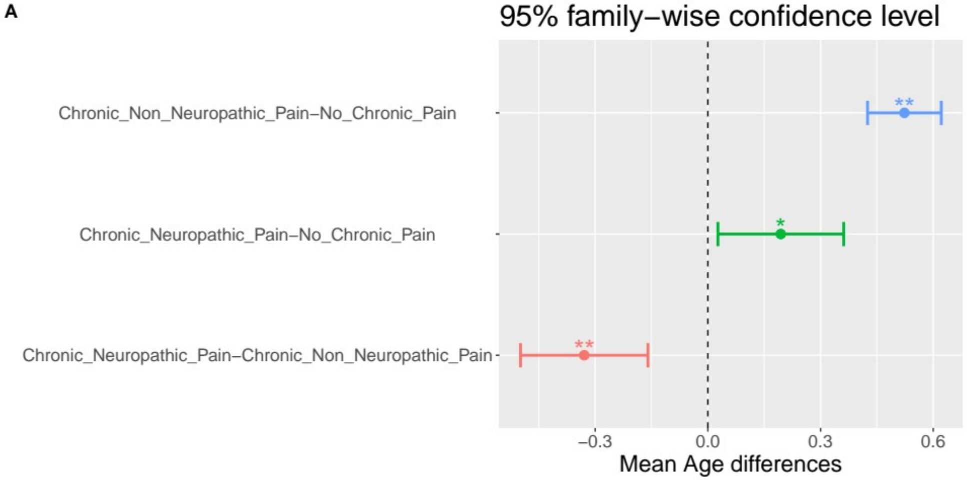


B

| | Dimension | Value | No_Chronic_Pain | Chronic_Non_Neuropathic_Pain | Chronic_Neuropathic_Pain |
|---|---|---|---|---|---|
| 1 | Female | Residuals | −26.0536308503014 | 21.0077953868443 | 9.19748932152672 |
| 2 | Female | p values | <0.0001 | <0.0001 | <0.0001 |
| 3 | Male | Residuals | 26.0536308503014 | −21.0077953868443 | −9.19748932152672 |
| 4 | Male | p values | <0.0001 | <0.0001 | <0.0001 |

C

| | Dimension | Value | No_Chronic_Pain | Chronic_Non_Neuropathic_Pain | Chronic_Neuropathic_Pain |
|---|---|---|---|---|---|
| 1 | BAME | Residuals | 5.19248137209141 | −6.25787700321982 | 1.702877 |
| 2 | BAME | p values | <0.0001 | <0.0001 | 0.797300 |
| 3 | Mixed | Residuals | −0.273187108541649 | −1.11060879532308 | 2.369724 |
| 4 | Mixed | p values | 1 | 1 | 0.160200 |
| 5 | White | Residuals | −4.34149950107226 | 5.97008443773198 | −2.683978 |
| 6 | White | p values | 1e−04 | <0.0001 | 0.065500 |

D

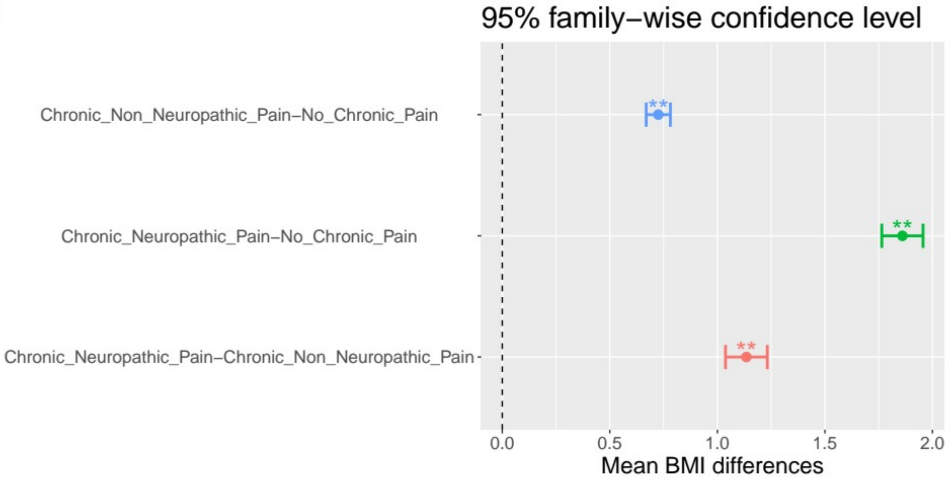


Supplementary Figure 1: Post-hoc follow-up tests for omnibus tests shown in figure 1. A: Tukey's HSD mean differences in age for all outcome levels. B-C: Chi-square post-hoc test for all levels of outcome. Standardised residuals and Bonferroni corrected p.values. D: Tukey's HSD mean differences in BMI for all outcome levels. P.value < 0.01 is coded as **, p.value < 0.05 is coded as *.

A

| | Dimension | Value | No_Chronic_Pain | Chronic_Non_Neuropathic_Pain | Chronic_Neuropathic_Pain |
|---|---|---|---|---|---|
| 1 | None of the above | Residuals | −0.327081121073469 | 0.250561684392678 | 0.137935074413102 |
| 2 | None of the above | p values | 1 | 1 | 1 |
| 3 | Prefer not to answer | Residuals | −0.206330431246903 | −0.165256411545335 | 0.638350660308744 |
| 4 | Prefer not to answer | p values | 1 | 1 | 1 |
| 5 | In paid employment or self−employed | Residuals | 11.6826503432546 | −8.57611719491137 | −5.56352762780595 |
| 6 | In paid employment or self−employed | p values | <0.0001 | <0.0001 | <0.0001 |
| 7 | Retired | Residuals | −8.00963341878631 | 8.29953370940934 | −0.311923644905685 |
| 8 | Retired | p values | <0.0001 | <0.0001 | 1 |
| 9 | Looking after home and/or family | Residuals | −0.250071215886216 | 0.49184451569524 | −0.406589959050507 |
| 10 | Looking after home and/or family | p values | 1 | 1 | 1 |
| 11 | Unable to work because of sickness or disability | Residuals | −18.1700576096873 | 2.16591721574574 | 27.7050269714519 |
| 12 | Unable to work because of sickness or disability | p values | <0.0001 | 0.8186 | <0.0001 |
| 13 | Unemployed | Residuals | −0.593032244873146 | −0.633739826311685 | 2.10546923052035 |
| 14 | Unemployed | p values | 1 | 1 | 0.9518 |
| 15 | Doing unpaid or voluntary work | Residuals | −0.0248588747275635 | 0.752431245317658 | −1.2401319209818 |
| 16 | Doing unpaid or voluntary work | p values | 1 | 1 | 1 |
| 17 | Full or part−time student | Residuals | −0.587864992638074 | 0.935158206195278 | −0.578834825035822 |
| 18 | Full or part−time student | p values | 1 | 1 | 1 |

B

| | Dimension | Value | No_Chronic_Pain | Chronic_Non_Neuropathic_Pain | Chronic_Neuropathic_Pain |
|---|---|---|---|---|---|
| 1 | Administrative and Secretarial Occupations | Residuals | −3.45542067652593 | 2.30006977 | 2.08487811030977 |
| 2 | Administrative and Secretarial Occupations | p values | 0.0165 | 0.64330000 | 1 |
| 3 | Associate Professional and Technical Occupations | Residuals | −5.53490219946127 | 3.68965473 | 3.33025434240247 |
| 4 | Associate Professional and Technical Occupations | p values | <0.0001 | 0.00670000 | 0.026 |
| 5 | Elementary Occupations | Residuals | −2.21409957396008 | −0.77697194 | 5.21835473971792 |
| 6 | Elementary Occupations | p values | 0.8047 | 1.00000000 | <0.0001 |
| 7 | Managers and Senior Officials | Residuals | 3.00323198538079 | −1.51558778 | −2.64603045083972 |
| 8 | Managers and Senior Officials | p values | 0.0801 | 1.00000000 | 0.2443 |
| 9 | Other job (free text entry) | Residuals | −1.9061375132423 | −0.31329329 | 3.87912224946455 |
| 10 | Other job (free text entry) | p values | 1 | 1.00000000 | 0.0031 |
| 11 | Personal Service Occupations | Residuals | −5.54841949824402 | 1.26530281 | 7.53579966711134 |
| 12 | Personal Service Occupations | p values | <0.0001 | 1.00000000 | <0.0001 |
| 13 | Process, Plant and Machine Operatives | Residuals | 0.0658633365120532 | −2.38637197 | 4.00099234342252 |
| 14 | Process, Plant and Machine Operatives | p values | 1 | 0.51050000 | 0.0019 |
| 15 | Professional Occupations | Residuals | 10.1660420174022 | −2.90477059 | −12.7958417781712 |
| 16 | Professional Occupations | p values | <0.0001 | 0.11030000 | <0.0001 |
| 17 | Sales and Customer Service Occupations | Residuals | −1.90989895159743 | −0.03016651 | 3.39733305480865 |
| 18 | Sales and Customer Service Occupations | p values | 1 | 1.00000000 | 0.0204 |
| 19 | Skilled Trades Occupations | Residuals | −1.76524280298689 | −0.12115468 | 3.30090893774506 |
| 20 | Skilled Trades Occupations | p values | 1 | 1.00000000 | 0.0289 |

C

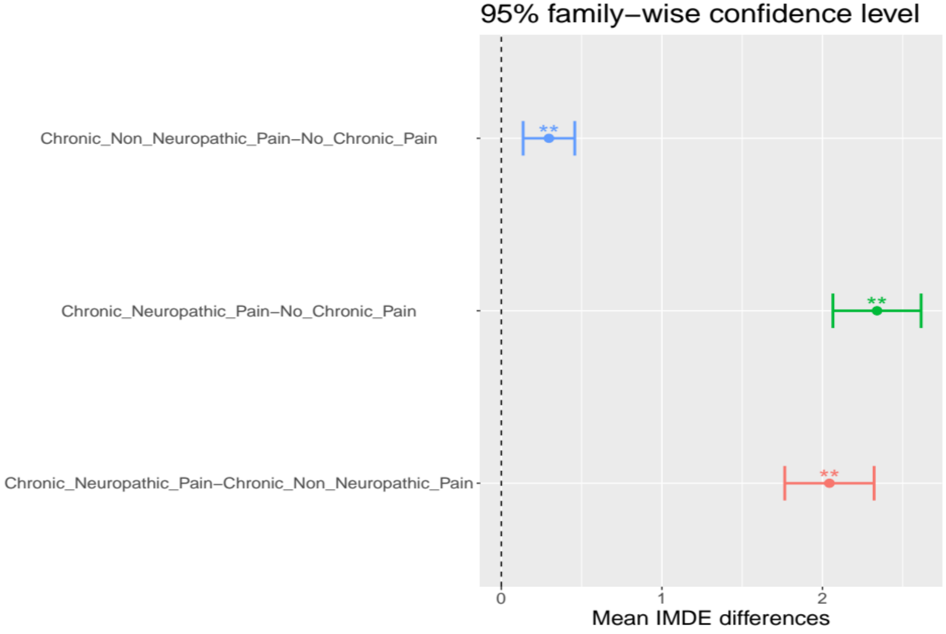

Supplementary Figure 2: Post-hoc follow-up tests for omnibus tests shown in figure 2. A-B:  Chi-square post-hoc test for all levels of outcome. Standardised residuals and Bonferroni corrected p.values. C: Tukey's HSD mean differences in Index of Multiple Deprivations England for all outcome levels. P.value < 0.01 is coded as **, p.value < 0.05 is coded as *.

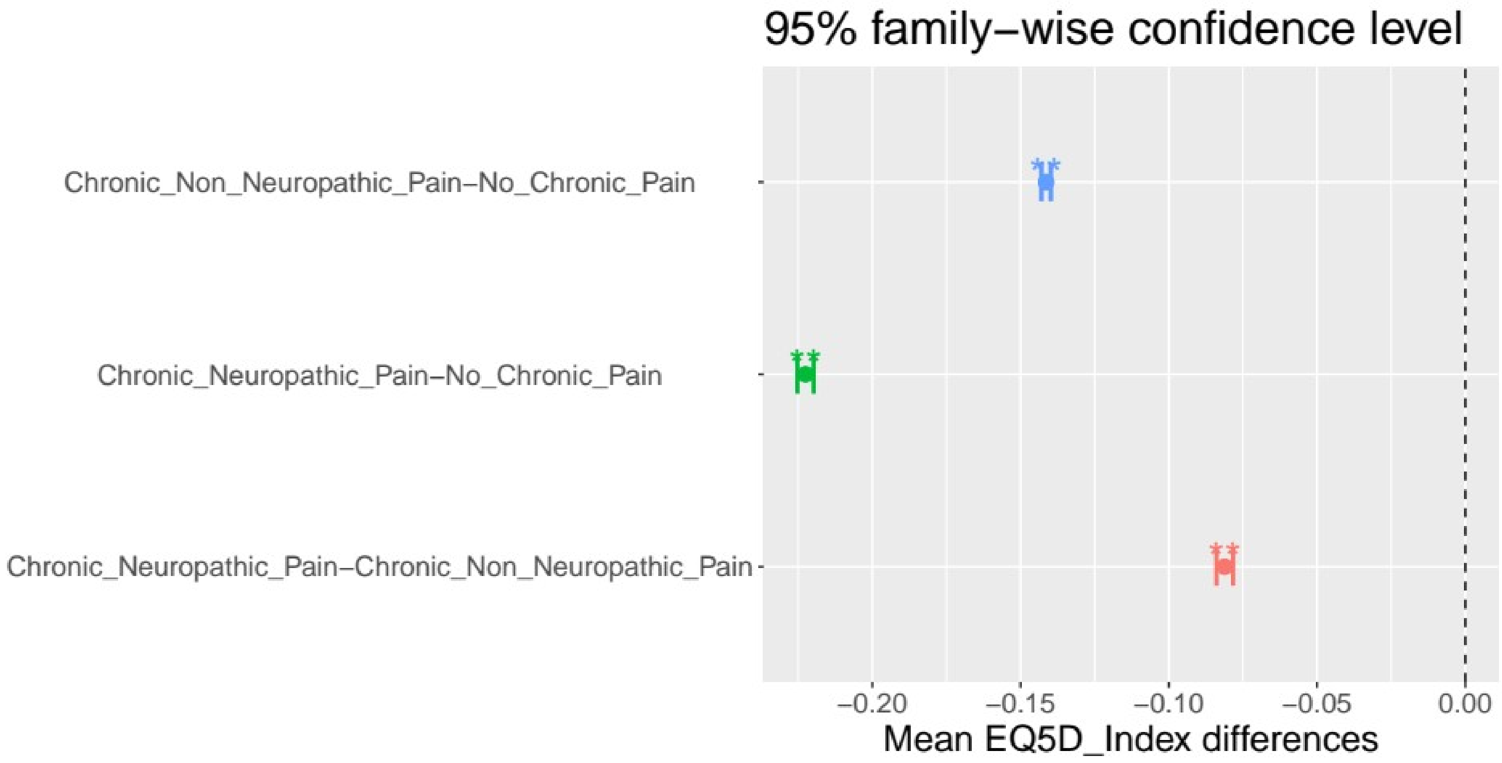


Supplementary Figure 3: Post-hoc follow-up tests for the omnibus test shown in figure 4. Tukey's HSD mean differences in EQ5D index for all outcome levels. P.value < 0.01 is coded as **, p.value < 0.05 is coded as *.

| | Dimension | Value | No_Chronic_Pain | Chronic_Non_Neuropathic_Pain | Chronic_Neuropathic_Pain |
|---|---|---|---|---|---|
| 1 | Certain infectious and parasitic diseases | Residuals | −6.26128993956141 | 1.67671090153365 | 7.59521869647907 |
| 2 | Certain infectious and parasitic diseases | p values | <0.0001 | 1 | <0.0001 |
| 3 | Congenital malformations, deformations and chromosomal abnormalities | Residuals | 0.854203757734353 | 0.215640249669986 | −1.76392271957834 |
| 4 | Congenital malformations, deformations and chromosomal abnormalities | p values | 1 | 1 | 1 |
| 5 | Diseases of the blood and blood−forming organs and certain disorders involving the immune mechanism | Residuals | −0.0117751667461538 | −1.73433637239848 | 2.85962513390601 |
| 6 | Diseases of the blood and blood−forming organs and certain disorders involving the immune mechanism | p values | 1 | 1 | 0.216 |
| 7 | Diseases of the circulatory system | Residuals | −2.23160060885637 | 2.39897374310772 | −0.242929210808806 |
| 8 | Diseases of the circulatory system | p values | 1 | 0.838 | 1 |
| 9 | Diseases of the digestive system | Residuals | −0.0887767003803269 | 0.608230576963175 | −0.849426383940015 |
| 10 | Diseases of the digestive system | p values | 1 | 1 | 1 |
| 11 | Diseases of the ear and mastoid process | Residuals | −0.49803087135157 | −0.266206435938577 | 1.25848234208916 |
| 12 | Diseases of the ear and mastoid process | p values | 1 | 1 | 1 |
| 13 | Diseases of the genitourinary system | Residuals | 2.44835766509529 | −0.251729055224606 | −3.63142550900219 |
| 14 | Diseases of the genitourinary system | p values | 0.732 | 1 | 0.014 |
| 15 | Diseases of the musculoskeletal system and connective tissue | Residuals | −1.53936695668768 | 2.45673815095091 | −1.48080512425075 |
| 16 | Diseases of the musculoskeletal system and connective tissue | p values | 1 | 0.715 | 1 |
| 17 | Diseases of the nervous system | Residuals | −17.2964855685302 | 6.75440468317291 | 17.5054245794863 |
| 18 | Diseases of the nervous system | p values | <0.0001 | <0.0001 | <0.0001 |
| 19 | Diseases of the respiratory system | Residuals | −0.0520245361272498 | −0.248451263677935 | 0.4927905738059 |
| 20 | Diseases of the respiratory system | p values | 1 | 1 | 1 |
| 21 | Diseases of the skin and subcutaneous tissue | Residuals | 2.46477858024095 | −2.1603603167067 | −0.532940340260565 |
| 22 | Diseases of the skin and subcutaneous tissue | p values | 0.699 | 1 | 1 |
| 23 | Endocrine, nutritional and metabolic diseases | Residuals | −3.37026705631578 | 2.1909145979522 | 1.97839175476566 |
| 24 | Endocrine, nutritional and metabolic diseases | p values | 0.038 | 1 | 1 |
| 25 | Factors influencing health status and contact with health services | Residuals | 0.884121663718701 | −0.160551217610325 | −1.1972772918993 |
| 26 | Factors influencing health status and contact with health services | p values | 1 | 1 | 1 |
| 27 | Injury, poisoning and certain other consequences of external causes | Residuals | 8.50516561561967 | −4.82427664223916 | −6.14665630590714 |
| 28 | Injury, poisoning and certain other consequences of external causes | p values | <0.0001 | <0.0001 | <0.0001 |
| 29 | Neoplasms | Residuals | 2.50555167594663 | −1.35025262766923 | −1.92692595857719 |
| 30 | Neoplasms | p values | 0.624 | 1 | 1 |
| 31 | Pregnancy, childbirth and the puerperium | Residuals | 9.22198767256997 | −4.92228977971558 | −7.1700381171355 |
| 32 | Pregnancy, childbirth and the puerperium | p values | <0.0001 | <0.0001 | <0.0001 |
| 33 | Symptoms, signs and abnormal clinical and laboratory findings, not elsewhere classified | Residuals | 5.67364989225804 | −2.6282614465453 | −5.06640886230518 |
| 34 | Symptoms, signs and abnormal clinical and laboratory findings, not elsewhere classified | p values | <0.0001 | 0.438 | <0.0001 |

Supplementary Figure 4: Chi-square post-hoc test for all levels of outcome. Standardised residuals and Bonferroni corrected p.values.
